# Future health gain from increasing physical activity in Australia, including multiple physiological effects of physical activity, and falls and injury risk: A simulation study

**DOI:** 10.64898/2026.03.28.26349629

**Authors:** Emily Bourke, Tim Wilson, Ralph Maddison, Tony Blakely

## Abstract

**Background:** Previous physical activity simulation studies only account for the effects of cardiovascular diseases, diabetes, dementia, and some cancers, which neglects many of its costs and benefits. We estimate the health and economic impacts of increased physical activity in Australia, including those on mental health, increased injury rate, and conditions mediated by other risk factors, commencing 2021, over 20 years.

**Methods:** We used a Proportional Multistate Lifetable Model specified with disease rate and risk factor forecasts, and causal associations, derived from the Global Burden of Disease study and other sources.

**Findings:** If all Australians shifted to the maximum physical activity level of 4200+ MET-min/week, there would be: 653,000 (230,000 - 1,210,000) or 0.16% more HALYs lived; 9,720 (7,400 to 12,700) or 1.33% fewer deaths before age 75; increased working age income of AUD$16.8 billion ($12.8 -$22.2 billion); and decreased health expenditure of $748 million (-$4.46 billion - $6.98 billion) or 0.02%.

Net health gains diminish for each additional 600 MET-min/week increase in physical activity, and above 4,200 MET-min/week the health costs from injuries outweigh the reduction in health costs from avoided disease. Because of injuries, increasing physical activity in the lowest activity group to meet the physical activity guidelines reduces health expenditure more ($1.86 billion; 896 million - 3.13 billion) than shifting to maximum activity levels.

**Interpretation:** Increasing physical activity levels in Australia would improve population health (even allowing for injuries due to participation), reduce health spending, and increase income.

**Funding:** Australian Sports Commission. TB is funded by NHMRC Investigator Grant (2023) #2026992

## Introduction

Physical activity can prevent many chronic diseases from occurring, including coronary heart disease, type 2 diabetes, stroke, dementia, and bowel, breast, and uterine cancers(1, 2). Recent work has expanded the range of conditions that are causally associated with physical inactivity. The Global Burden of Disease (GBD) Study 2021(1) now includes the flow-on effects from diabetes caused by physical inactivity, such as chronic kidney disease. However, further modelling has expanded the physical inactivity model to estimate the impact of physical activity onto depression and anxiety, and through other risk factors not captured in the GBD study(3), including high blood pressure, high fasting plasma glucose, high LDL-cholesterol levels, and low bone mineral density, and found that these additions increase attributable burden by about 60%.

On the other hand, injuries can also be sustained during physical activity participation (such as cycling accidents, or fractures from playing sports) (4), which may reduce the beneficial effects of physical activity. Some studies have included the effect of active transport related injuries in assessments of transport interventions(5, 6), such as increased injuries for cyclists when the transport mode is shifted from driving to cycling. A systematic review of such transport intervention studies (n=30) found that the estimated benefits to health in terms of diseases prevented, and the estimated risks or costs to health due to increased injuries caused by activity, had a median ratio of 9:1 of the benefits to costs/risks from increasing physical activity(7). However, the trade-off of increasing physical activity participation in terms of diseases prevented and injuries sustained during activity has not been assessed for total physical activity levels.

This study aimed to estimate the future changes over time in the health and economic impacts of physical activity in Australia under various participation scenarios. This study is the first to model both the health gains of expanding physical activity benefits to include all conditions caused by risk factors impacted by physical activity, and the offsetting health loss due to injuries sustained while participating in physical activity, to generate ‘net’ health gains. We included as diseases and conditions causally effected by physical inactivity: a) those associated with physical inactivity in the GBD and Australian Burden of Disease Study (coronary heart disease, type 2 diabetes, stroke, dementia, and bowel, breast, and uterine cancers); b) depression and anxiety; c) diseases due to the causal effect of physical activity on the intermediate risk factors high blood pressure, fasting plasma glucose (and type 2 diabetes) and LDL-cholesterol, and low bone mineral density; and d) the risk of injuries from undertaking physical activity.

We used a Proportional Multistate Lifetable Model to simulate the Australian population over 20 years to calculate the differences in health adjusted life years (HALYs), years of life, health expenditure, income loss, and deaths between the forecast business-as-usual, and four intervention scenarios ranging from highest to lowest total activity participation: whole population achieving highest level of physical activity, those not meeting guidelines increasing activity to meet guidelines, population reverting to earlier (2018) lower levels of physical activity, the whole population becoming sedentary, and iteratively shifting the population to the next highest physical activity group.

These five scenarios enable us to answer three key research questions: 1) what is the benefit gained from the current level of physical activity undertaken (compared to everyone inactive), and how much more health improvements can be made by increasing activity levels, 2) what is the negative effect of injuries compared to the positive effect on disease risk, and 3) at what level of physical activity does the injury negative effect outweigh the positive disease risk reductions? Results are tested under varying time horizons and discount rates.

## Methods

We adapted a proportional multistate lifetable model (Blakely et al 2020 (8)) for this paper. The PMSLT has been expanded using the risk factor pathways and directly linked conditions described in Bourke et al 2026(3) to estimate both directly and indirectly the disease risk from physical inactivity, and including incident injuries caused by participation in physical activity. We used the physical activity distribution of the population, and the estimated distribution of blood pressure, fasting plasma glucose, LDL-cholesterol, and bone mineral density within each level of physical activity for each age and sex group, to model the changes to disease burden (health adjusted life years (HALYs) and deaths), health spending, and income loss for different scenarios of activity participation.

### Data inputs

Table 1 outlines the data sources and inputs used for each component of the model. Details on the derivation of the dose response of physical activity onto other mediating risk factors, estimating distributions of each risk factor by level of physical activity, calculation of disease phase transition rates, rate ratios of disease and injury risk by level of physical activity, and details of costing data are included in the Supplementary Material.

**Table 1.** data sources and inputs. *Note: ABS = Australian Bureau of Statistics, GBD = Global Burden of Disease, YLD = Years Lived With Disease, LDL = Low density lipoprotein, MET = Metabolic Equivalent of Task, RF = risk factor, AIHW = Australian Institute of Health and Welfare, ABDS = Australian Burden of Disease Study*

| Model component | Input | Source |
| --- | --- | --- |
| **Underlying population characteristics** | Australian Bureau of Statistics population estimates by age and sex for 2021 | ABS 2021(9) |
|  | Birth projections by sex | ABS 2021(9) |
|  | Incidence, prevalence, remission, case fatality rates by age and sex for all diseases and injuries | GBD 2021 (1) |
|  | Disability rates for each condition (i.e. YLDs for that condition by sex and age, divided by the number of people in that sex and age group) | GBD 2021 (1) |
| **Risk factor distributions** | Australian population distributions by age and sex of the risk factors physical activity, blood pressure, blood glucose, LDL-cholesterol, and bone mineral density<br>Physical activity: percent of population in 600 MET-min/week increments to 4200+<br>Other RF distributions: calculated from mean and standard deviation of each risk factor | Australian Burden of Disease Study 2024(2) |
|  | Continuous variable distribution of fasting plasma glucose, systolic blood pressure, and bone mineral density by physical activity level, derived from relative risks of diabetes, hypertension, and osteoporosis dichotomies with physical activity. See Supplementary Table S3 for details. | Kyu et al 2016(10), Liu et al 2017(11), Pinheiro et al 2020 (12) |
|  | Distribution of continuous LDL-cholesterol by physical activity level from analysis of UK Biobank data. See Supplementary Table S3 for details. | UK Biobank(13) |
| **Risk of conditions and injuries** | Incidence rate ratios from each risk factor (physical activity, blood pressure, fasting plasma glucose, diabetes, and LDL-cholesterol; by age, sex, and risk factor exposure level), for each of the conditions currently linked to each risk factor (i.e. accepted risk outcome pairs). See Supplementary Table S3 for details. | GBD 2019(14) |
|  | Estimate of relative risk of physical inactivity for depression and anxiety incidence | Schuch et al, 2019(15), Schuch et al, 2018(16) |
|  | The proportion of all injuries estimated to be due to physical activity for each sex and age group was taken from previous AIHW work (percent of hospitalised injuries identified as due to activity, by age, sex, and injury type. See Supplementary Table S2 for percentages.). We then assumed a pro-rata or linear association of MET-min/week of activity with injury incidence and generated an injury rate per MET-min/week, by age and sex. | AIHW 2024(4, 17), GBD 2021(1), ABDS 2024(2). |
| **Disease costs** | Total healthcare cost of each disease in a year in Australia for each age and sex | AIHW 2021(18) |
|  | Relative cost of each disease by phase of disease (incident year, last year of life, and otherwise prevalent). | Blakely et al, 2019(19) |
|  | Estimated loss of income due to disease diagnosis | Blakely et al, 2021(20) |

### PMSLT

The PMSLT methodology is described in detail elsewhere(8). The model base year is 2021 and uses a main cohort lifetable to simulate the Australian population as an open cohort (with annual estimated births and net migration), with 15-year trend projections of all-cause morbidity and mortality rates and cause-specific disease rates (above) by age and sex – then rates held constant. The default time horizon was 20-years for reporting outputs of health adjusted life years (HALYs), net changes in health expenditure and income – but we also plotted outputs by year over 40 years to allow inspection of variation by time. The results are undiscounted (3% reported in sensitivity analyses), and monetary estimates are reported in 2023 AUD.

The ‘business as usual’ (BAU) scenario was the proportion of each age and sex group in each physical activity level according to baseline exposure. We assumed no change over time in the physical activity distribution, though cohorts change to the next age group’s physical activity and risk factor exposure level as they age.

No time lags were modelled for a change in physical activity to a change in the other risk factors (i.e. causation instantaneous). Change in disease risk from physical activity directly and all intermediate risk factors had variable time lags applied (refer to Supplementary Table 5 for each conditions time lag). Time lags were implemented with a look back period of 5 to 20 years in the past (for short and long duration conditions) to determine the weighted average risk for each year. Monte Carlo simulation was used to estimate uncertainty with +/- 20% on the ln-normal scale.

Differences in each of the model outputs (HALYs, life years, deaths, income, healthcare spending) between the BAU scenario and each of the intervention scenarios were calculated for each year following the base year and represented the impact of the change in physical activity level.

The analysis was conducted in Python, and code is not publicly available.

### Intervention scenarios

The intervention scenarios modelled were theoretical distributions of physical activity, rather than specific interventions to try and increase physical activity. Some scenarios reflected an increase in physical activity, to estimate how much gains can be made to health, and some reflected reductions in physical activity, to estimate what the benefit of the current level of physical activity was. The comparator business-as-usual scenario is 2022 levels of physical activity, by sex and period age, continuing into the future.

The scenarios, in order of highest to lowest total activity undertaken, were: 1) **Maximal activity**: 100% of each age and sex reached the highest physical activity category (4200+ MET-min/week), 2) **Inactivity eliminated**: everyone in the 0-600 MET-min/week group moved to the 600-1200 MET-min/week group (meeting minimum total physical activity recommendations), 3) **Revert to 2018**: the population reverting to the lesser 2018 physical activity level for each age and sex, 4) **Everyone inactive**: all sex by age groups set to 0-600 MET-min/week, and 5) **Thresholds**: To estimate the differences in effect of physical activity for each activity level threshold, the population in each activity level is shifted to the next highest (until the 3600-4200 MET-min/week group reaches 4200+). The change for each group was modelled independently of the changes to the other groups. In all scenarios, relative risks for diseases were applied to the population aged 20 and over.

### Sensitivity analyses

#### Discount rate

main model outputs were run with a standard 3% discount rate.

#### Removing additional diseases and conditions

The main model included depression and anxiety, conditions impacted by risk factors that physical activity impacts, and offsetting health loss from injury due to increasing physical activity. To allow comparison with previous research, we reran the model excluding these extensions. The main model including all pathways was Model 1. Model 2 included all the additional direct conditions and risk factor pathways to disease but excludes physical activity causing incident injuries. Model 3 reflected only the conditions that were included in standard models of physical activity (coronary heart disease, stroke, type 2 diabetes, bowel cancer, breast cancer, uterine cancer, and dementia(2)), and excluded any pathway to account for overadjustment of relative risks.

#### No time lags

Changes in relative risks for all conditions have no time lags applied (model 4).

#### Alternate method of distributing injuries with physical activity

Injury incidence was distributed by total MET-min/week in the main model, which captured both intensity of activity and total time spent active. An alternate method to distribute injury incidence by activity was tested by using only the total time undertaking activity, rather than MET-min/week (model 5). We assumed that the average intensity of activity in the 0-600 MET-min/week group was 4 METs, increasing by 0.5 METs in each group to 7.5 MET in the 4200+ group.

## Results

Table 2 presents the undiscounted health and economic impacts of scenarios 1 to 4, compared to BAU, over a 20-year time horizon. Figure 1 presents the same metrics, for each year over a 40-year time horizon to allow determination of time varying impacts. Table 3 reports the contribution of condition groups to overall changes in HALY and disease expenditure savings for scenarios 1 and 4. Figure 2 reports the results of the threshold analysis for scenarios 5. Table 4 includes sensitivity analysis results and explores the impact of including a wider number of diseases and injuries in the base model in this paper, compared to a reduced list of conditions as per previous modelling.

**Table 2.** Total HALYS, health spending, income, and deaths over 20 years in BAU, and the difference for scenarios 1 to 4 compared to BAU, by sex, 0% discount rate. Note: 1) Maximal activity: 100% of each age and sex reached the highest physical activity category (4200+ MET-min/week), 2) Meet guidelines: everyone in the 0-600 MET-min/week group moved to the 600-1200 MET-min/week group (meeting minimum total physical activity guidelines), 3) Revert to 2018: the population reverting to the lesser 2018 physical activity level for each age and sex, 4) Everyone inactive: all sex by age groups set to 0-600 MET-min/week

| Metric | Sex | Scenarios 1 to 4 compared to BAU (current physical activity level) |  |  |  |  |
| --- | --- | --- | --- | --- | --- | --- |
|  |  | BAU | 1. Maximal activity | 2. Meet guidelines | 3. Revert to 2018 | 4. Everyone inactive |
| Total HALY (thousands) | All | 398,000 | 653 (230 to 1,210) | 213 (131 to 328) | -151 (-244 to -84.3) | -1,290 (-2,160 to -679) |
|  | Female | 199,000 | 450 (192 to 790) | 119 (72.7 to 185) | -92.3 (-150 to -51.6) | -684 (-1,130 to -378) |
|  | Male | 199,000 | 202 (37 to 418) | 93.7 (57.8 to 143) | -58.5 (-94.7 to -32.5) | -608 (-1,030 to -307) |
| Life years gained (thousands) | All | 475,000 | 258 (193 to 343) | 41.2 (23.7 to 63.4) | -32.2 (-45.1 to -22.8) | -221 (-324 to -144) |
|  | Female | 242,000 | 129 (96.1 to 171) | 20.8 (11.7 to 32.4) | -16.9 (-23.7 to -11.8) | -85.9 (-125 to -56.1) |
|  | Male | 234,000 | 129 (96.5 to 171) | 20.3 (11.9 to 31.2) | -15.3 (-21.5 to -10.9) | -135 (-199 to -88.4) |
| Total health spending (millions, AUD) | All | 3,580,000 | -748 (-6,980 to 4,460) | -1,860 (-3,130 to -896) | 1,450 (655 to 2,490) | 9,800 (3,320 to 18,600) |
|  | Female | 1,880,000 | -2,060 (-5,900 to 1,010) | -1,020 (-1,740 to -484) | 819 (358 to 1,430) | 5,860 (2,710 to 10,300) |
|  | Male | 1,700,000 | 1,330 (-1,310 to 3,510) | -845 (-1,410 to -395) | 625 (276 to 1,080) | 3,960 (566 to 8,470) |
| Total income (millions, AUD) | All | 11,800,000 | 16,800 (12,800 to 22,200) | 1,100 (705 to 1,660) | -1,970 (-2,670 to -1,450) | -16,900 (-23,200 to -12,000) |
|  | Female | 4,710,000 | 9,030 (6,870 to 12,000) | 500 (328 to 736) | -1,060 (-1,430 to -789) | -6,820 (-9,380 to -4,890) |
|  | Male | 7,060,000 | 7,800 (5,910 to 10,200) | 606 (363 to 962) | -909 (-1,240 to -650) | -10,100 (-14,000 to -6,980) |
| Total deaths | All | 4,300,000 | -33,700 (-43,700 to -25,700) | -4,730 (-7,240 to -2,770) | 4,420 (3,170 to 6,060) | 28,700 (19,100 to 41,100) |
|  | Female | 2,070,000 | -16,800 (-21,900 to -12,800) | -2,380 (-3,660 to -1,370) | 2,250 (1,620 to 3,070) | 11,000 (7,430 to 15,700) |
|  | Male | 2,220,000 | -16,800 (-21,900 to -12,800) | -2,350 (-3,610 to -1,390) | 2,170 (1,550 to 2,980) | 17,700 (11,800 to 25,600) |
| Deaths before age 75 | All | 1,010,000 | -9,720 (-12,700 to -7,400) | -931 (-1,540 to -535) | 1,590 (1,140 to 2,230) | 12,100 (8,180 to 17,900) |
|  | Female | 392,000 | -3,550 (-4,550 to -2,750) | -284 (-453 to -168) | 557 (409 to 758) | 3,200 (2,220 to 4,570) |
|  | Male | 621,000 | -6,150 (-8,210 to -4,610) | -647 (-1,090 to -358) | 1,040 (727 to 1,470) | 8,930 (5,860 to 13,300) |

**Table 3.** Disease groups contribution to total difference in HALYs and health cost, over 20 years from the BAU in scenarios 1 to 4, over 20 years with 0% discount rate.

| Metric and Condition | 1. Maximal activity |  |  | 2. Meet guidelines |  |  | 3. Revert to 2018 |  |  | 4. Everyone inactive |  |  |
| --- | --- | --- | --- | --- | --- | --- | --- | --- | --- | --- | --- | --- |
|  | All | Female | Male | All | Female | Male | All | Female | Male | All | Female | Male |
| **Total HALY (thousands)** | **653** | **450** | **202** | **213** | **119** | **93.7** | **-151** | **-92.3** | **-58.5** | **-1,290** | **-684** | **-608** |
| **(95% UI)** | **(230 to 1,210)** | **(192 to 790)** | **(37 to 418)** | **(131 to 328)** | **(72.7 to 185)** | **(57.8 to 143)** | **(-244 to -84.3)** | **(-150 to -51.6)** | **(-94.7 to -32.5)** | **(-2,160 to -679)** | **(-1,130 to -378)** | **(-1,030 to -307)** |
| **Cancer and other neoplasms** | 44.5 | 26.2 | 18.5 | 2.41 | 1.63 | 0.8 | -5.98 | -3.56 | -2.41 | -31.7 | -16.4 | -15.4 |
| **(95% UI)** | **(25 to 75.1)** | **(15.3 to 41.7)** | **(8.7 to 33.8)** | **(-0.51 to 6.99)** | **(-0.03 to 4.05)** | **(-0.58 to 3.02)** | **(-10.8 to -2.97)** | **(-6.16 to -1.9)** | **(-4.73 to -1.01)** | **(-62.8 to -13.4)** | **(-29.9 to -7.6)** | **(-33.2 to -4.99)** |
| **Cardiovascular diseases** | 191 | 90.5 | 100 | 32.9 | 15.7 | 17.2 | -25.3 | -12.4 | -12.9 | -183 | -65.9 | -117 |
| **(95% UI)** | **(135 to 260)** | **(64.3 to 124)** | **100 (71.1 to 137)** | **(19.0 to 51.4)** | **(8.74 to 24.5)** | **(10.2 to 26.8)** | **(-36.7 to -16.8)** | **(-17.9 to -8.18)** | **(-18.9 to -8.62)** | **(-276 to -116)** | **(-99.5 to -40.8)** | **(-177 to -74.3)** |
| **Endocrine disorders** | 239 | 116 | 122 | 5.08 | 2.5 | 2.59 | -27.1 | -13.2 | -13.9 | -161 | -59.9 | -101 |
| **(95% UI)** | **(171 to 329)** | **(82.9 to 161)** | **(88.2 to 168)** | **(1.3 to 10.3)** | **(0.55 to 5.06)** | **(0.58 to 5.26)** | **(-40.1 to -17.2)** | **(-19.4 to -8.55)** | **(-20.8 to -8.66)** | **(-252 to -93.7)** | **(-95.4 to -33.9)** | **(-157 to -59.8)** |
| **Hearing and vision disorders** | 0.44 | 0.23 | 0.2 | 0.009 | 0.005 | 0.003 | -0.04 | -0.02 | -0.02 | -0.27 | -0.11 | -0.16 |
| **(95% UI)** | **(0.03 to 1.05)** | **(0.02 to 0.57)** | **(0.01 to 0.48)** | **(0.0 to 0.03)** | **(0.0 to 0.02)** | **(0.0 to 0.01)** | **(-0.01 to -0.002)** | **(-0.06 to -0.002)** | **(-0.05 to -0.001)** | **(-0.69 to -0.02)** | **(-0.29 to -0.007)** | **(-0.41 to -0.001)** |
| **Injuries** | -761 | -360 | -400 | -0.21 | 0.54 | -0.77 | 54.1 | 27.6 | 26.5 | 421 | 146 | 276 |
| **(95% UI)** | **(-884 to -654)** | **-360 (-426 to -302)** | **(-458 to -352)** | **(-17 to 17.4)** | **(-8.05 to 9.7)** | **(-8.96 to 7.86)** | **(43.4 to 65.7)** | **(21.3 to 34.2)** | **(22.1 to 31.3)** | **(344 to 507)** | **(111 to 182)** | **(233 to 325)** |
| **Kidney and urinary diseases** | 14.5 | 6.8 | 7.66 | 0.54 | 0.27 | 0.27 | -1.45 (-2.45 to -0.78) | -0.72 | -0.74 | -7.48 | -2.89 | -4.59 |
| **(95% UI)** | **(7.54 to 25.7)** | **(3.58 to 12.1)** | **(4.01 to 13.6)** | **(0.27 to 1.0)** | **(0.13 to 0.49)** | **(0.14 to 0.51)** | **(-2.45 to -0.78)** | **(-1.23 to -0.39)** | **(-1.23 to -0.4)** | **(-13 to -3.96)** | **(-5.02 to -1.53)** | **(-7.92 to -2.43)** |
| **Mental and substance use disorders** | 866 | 534 | 333 | 168 | 95.7 | 72.4 | -139 | -85.9 | -53.1 | -1,300 | -668 | -631 |
| **(95% UI)** | **(485 to 1,390)** | **(298 to 861)** | **(186 to 531)** | **(89 to 279)** | **(50.8 to 160)** | **(38.5 to 119)** | **(-227 to -75.2)** | **(-141 to -46.7)** | **(-86.1 to -28.9)** | **(-2,130 to -700)** | **(-1,100 to -359)** | **(-1,030 to -341)** |
| **Neurological conditions** | 44.5 | 28.5 | 16 | 2.83 | 1.97 | 0.87 | -5.07 | -3.44 | -1.64 | -23.8 | -14 | -9.8 |
| **(95% UI)** | **(10.5 to 98.5)** | **(6.44 to 63.6)** | **16 (4 to 34.8)** | **(0.36 to 7.11)** | **(0.23 to 4.98)** | **(0.13 to 2.12)** | **(-11.4 to -1.08)** | **(-7.82 to -0.69)** | **(-3.57 to -0.37)** | **(-54.8 to -4.75)** | **(-32.7 to -2.61)** | **(-22.1 to -2.1)** |

| **Total health spending**<br>**(millions, AUD)**<br>**(95% UI)** | **-748**<br>**(-6,980 to**<br>**4,460)** | **-2,060**<br>**(-5,900 to**<br>**1,010)** | **1,330**<br>**(-1,310 to**<br>**3,510)** | **-1,860**<br>**(-3,130 to -**<br>**896)** | **-1,020**<br>**(-1,740 to**<br>**-484)** | **-845**<br>**(-1,410**<br>**to -395)** | **1,450**<br>**(655 to**<br>**2,490)** | **819**<br>**(358 to**<br>**1,430)** | **625**<br>**(276 to 1,080)** | **9,800**<br>**(3,320 to**<br>**18,600)** | **5,860**<br>**(2,710 to**<br>**10,300)** | **3,960**<br>**(566 to**<br>**8,470)** |
| --- | --- | --- | --- | --- | --- | --- | --- | --- | --- | --- | --- | --- |
| **Cancer and other**<br>**neoplasms**<br>**(95% UI)** | -1,310<br>(-2,200 to -<br>720) | -862<br>(-1,420 to<br>-491) | -449<br>(-844 to -<br>202) | -52.7<br>(-163 to 4.45) | -42.1<br>(-119 to<br>0.76) | -9.7<br>(-53.1 to<br>8.97) | 183<br>(85.7 to<br>345) | 125<br>(58.7 to<br>222) | 58.4<br>(19.5 to 126) | 1,100<br>(457 to<br>2,150) | 674<br>(265 to<br>1,290) | 417<br>(121 to 955) |
| **Cardiovascular diseases**<br>**(95% UI)** | -2,350<br>(-4,160 to<br>-1,110) | -726<br>(-1,420 to<br>-198) | -1,620<br>(-2,750 to<br>-855) | -40.1<br>(-385 to 171) | 49.3<br>(-63.5 to<br>161) | -94.1<br>(-331 to<br>32.3) | 514<br>(276 to<br>922) | 142<br>(59.6 to<br>279) | 375<br>(212 to 642) | 3,510<br>(1,670 to<br>6,800) | 742<br>(282 to<br>1,610) | 2,770<br>(1,360 to<br>5,190) |
| **Endocrine disorders**<br>**(95% UI)** | -4,400<br>(-6,530 to<br>-2,990) | -2,010<br>(-2,990 to -<br>1,360) | -2,390<br>(-3,540 to -<br>1,630) | -90.7<br>(-186 to -<br>20.4) | -41.6<br>(-85.6 to -<br>9.39) | -49.1<br>(-100 to -<br>11.1) | 496<br>(305 to<br>783) | 224<br>(140 to<br>353) | 272<br>(165 to 431) | 3,050<br>(1,720 to<br>4,990) | 1,050<br>(573 to<br>1,750) | 2,000<br>(1,140 to<br>3,250) |
| **Injuries**<br>**(95% UI)** | 16,900<br>(15,100 to<br>18,600) | 7,230<br>(6,120 to<br>8,230) | 9,690<br>(8,930 to<br>10,400) | 61.9<br>(-307 to 383) | -18.1<br>(-222 to<br>162) | 80.7<br>(-84.8 to<br>220) | -1,240<br>(-1,420 to<br>-1,050) | -561<br>(-673 to -<br>432) | -685<br>(-743 to -619) | -10,200<br>(-11,500<br>to -<br>8,840) | -2,880<br>(-3,470 to<br>-2,180) | -7,360<br>(-8,000 to -<br>6,640) |
| **Kidney and urinary**<br>**diseases**<br>**(95% UI)** | -714<br>(-1,250 to<br>-373) | -281<br>(-493 to -<br>147) | -433<br>(-753 to -<br>227) | -22<br>(-41.1 to -<br>10.7) | -9.03<br>(-16.9 to -<br>4.38) | -13<br>(-24.1 to<br>-6.36) | 81.8<br>(43.8 to<br>140) | 32.1<br>(17 to 55.7) | 49.7<br>(26.8 to 85) | 468<br>(248 to<br>821) | 146<br>(76.4 to<br>257) | 323<br>(171 to 563) |
| **Mental and substance**<br>**use disorders**<br>**(95% UI)** | -7,960<br>(-13,500 to<br>-4,270) | -4,970<br>(-8,440 to<br>-2,640) | -2,980<br>(-5,060 to<br>-1,610) | -1,640<br>(-2,860 to -<br>851) | -925<br>(-1,620 to<br>-473) | -716<br>(-1,250<br>to -372) | 1,300<br>(679 to<br>2,210) | 800<br>(418 to<br>1,380) | 492<br>(262 to 844) | 11,200<br>(5,930 to<br>19,200) | 5,880<br>(3,070 to<br>10,100) | 5,320<br>(2,830 to<br>9,130) |
| **Neurological conditions**<br>**(95% UI)** | -580<br>(-1,330 to -<br>133) | -284<br>(-660 to -<br>61.4) | -296<br>(-672 to -<br>71.5) | -36.7<br>(-95.1 to -<br>4.66) | -20.4<br>(-53.5 to -<br>2.43) | -16.3<br>(-41.5 to<br>-2.41) | 65.8<br>(13.3 to<br>152) | 34.6<br>(6.44 to<br>81.9) | 31.1<br>(6.92 to 70.3) | 317<br>(59.9 to<br>748) | 137<br>(23.5 to<br>331) | 179<br>(36.4 to<br>416) |

**Table 4.** Sensitivity analysis results for scenario 1 (maximal activity) with disease risk scenarios including all pathways, all pathways excluding injury, only standard conditions, and alternate distribution for injuries by physical activity level, over 20 years, 0% discount rate.

| Metric | Sex | 1. All pathways including injury (main analysis) | 2. All pathways, no injury | 3. Standard conditions only | 4. No lags | 5. Injuries by time, not METS |
| --- | --- | --- | --- | --- | --- | --- |
| **Total HALY (thousands)** | All | 653 (230 to 1,210) | 1,520 (1,100 to 2,060) | 425 (333 to 551) | 832 (385 to 1,370) | 650 (213 to 1,220) |
|  | Female | 450 (192 to 790) | 874 (616 to 1,210) | 218 (169 to 288) | 544 (276 to 883) | 450 (183 to 804) |
|  | Male | 202 (37 to 418) | 647 (487 to 860) | 206 (161 to 265) | 286 (102 to 505) | 203 (23.8 to 414) |
| **Total health spending (millions, AUD)** | All | -748 (-6,980 to 4,460) | -19,900 (-25,800 to -15,300) | -6,810 (-9,200 to -4,740) | -3,140 (-9,180 to 2,120) | -607 (-7,450 to 4,490) |
|  | Female | -2,060 (-5,900 to 1,010) | -10,700 (-14,500 to -8,040) | -3,200 (-4,330 to -2,190) | -3,280 (-6,860 to -299) | -2,000 (-6,050 to 897) |
|  | Male | 1,330 (-1,310 to 3,510) | -9,150 (-11,600 to -7,130) | -3,600 (-4,930 to -2,510) | 112 (-2,560 to 2,480) | 1,380 (-1,440 to 3,530) |
| **Total income (millions)** | All | 16,800 (12,800 to 22,200) | 17,600 (13,800 to 22,500) | 13,000 (9,720 to 17,900) | 20,700 (16,400 to 26,400) | 16,800 (13,000 to 21,700) |
|  | Female | 9,030 (6,870 to 12,000) | 9,200 (7,170 to 11,900) | 6,830 (5,020 to 9,580) | 11,100 (8,720 to 14,000) | 9,020 (6,950 to 11,500) |
|  | Male | 7,800 (5,910 to 10,200) | 8,340 (6,610 to 10,500) | 6,180 (4,670 to 8,380) | 9,610 (7,600 to 12,300) | 7,840 (5,970 to 10,200) |
| **Total deaths** | All | -33,700 (-43,700 to -25,700) | -33,500 (-43,500 to -25,700) | -22,200 (-30,500 to -16,200) | -42,900 (-54,400 to -34,600) | -34,000 (-44,700 to -26,500) |
|  | Female | -16,800 (-21,900 to -12,800) | -16,700 (-21,600 to -12,900) | -11,300 (-15,500 to -8,260) | -21,600 (-27,100 to -17,300) | -17,000 (-22,400 to -13,200) |
|  | Male | -16,800 (-21,900 to -12,800) | -16,800 (-21,900 to -12,700) | -10,900 (-15,100 to -7,850) | -21,400 (-27,400 to -17,200) | -16,900 (-22,300 to -13,200) |

**Figure 1.**
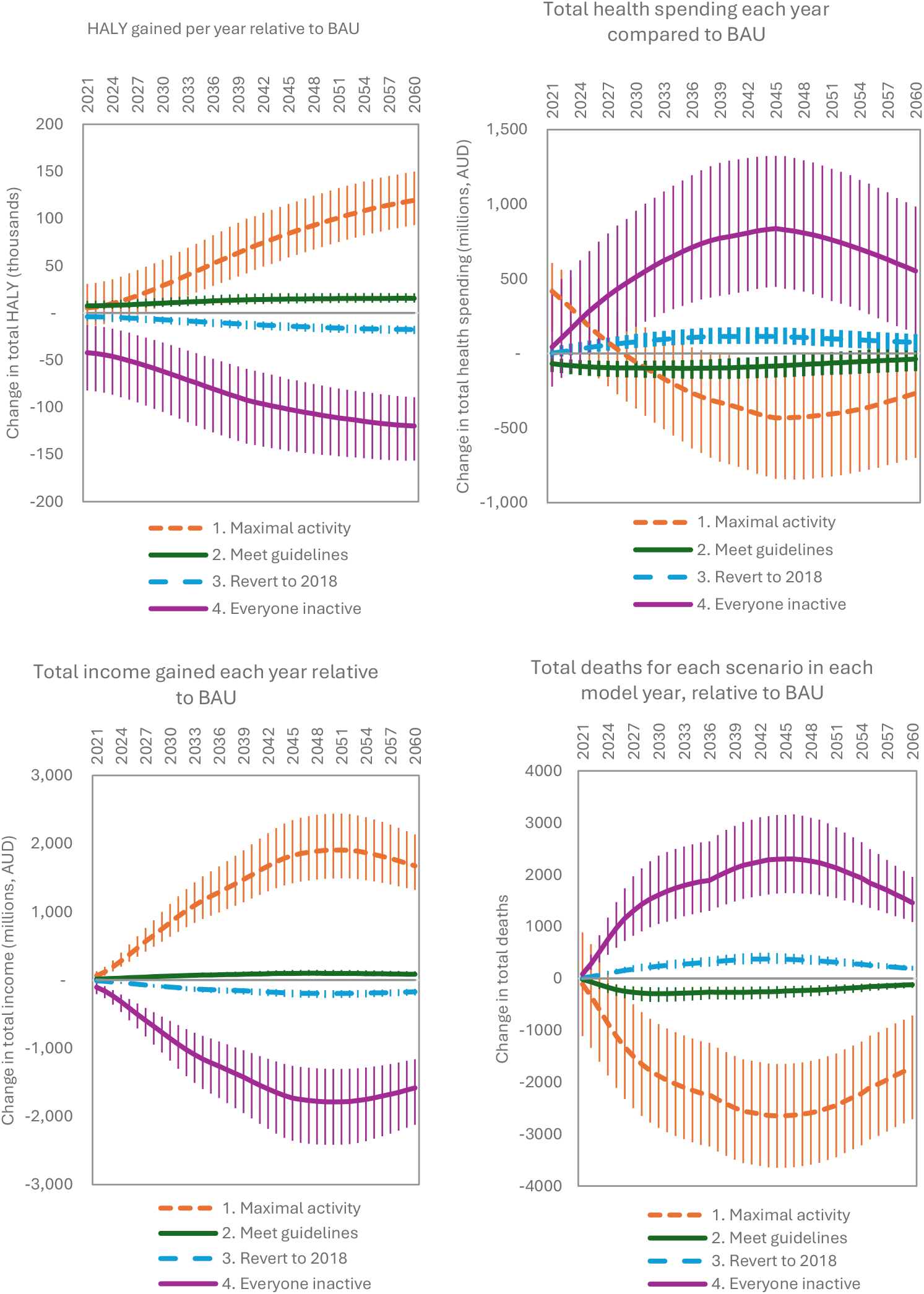
Total HALYS, health spending, income, and deaths, relative to BAU for 4 main intervention scenarios, each year for 40 years with 0% discount rate. Note: 1) Maximal activity: 100% of each age and sex reached the highest physical activity category (4200+ MET-min/week), 2) Meet guidelines: everyone in the 0-600 MET-min/week group moved to the 600-1200 MET-min/week group (meeting minimum total physical activity guidelines), 3) Revert to 2018: the population reverting to the lesser 2018 physical activity level for each age and sex, 4) Everyone inactive: all sex by age groups set to 0-600 MET-min/week

**Figure 2.**
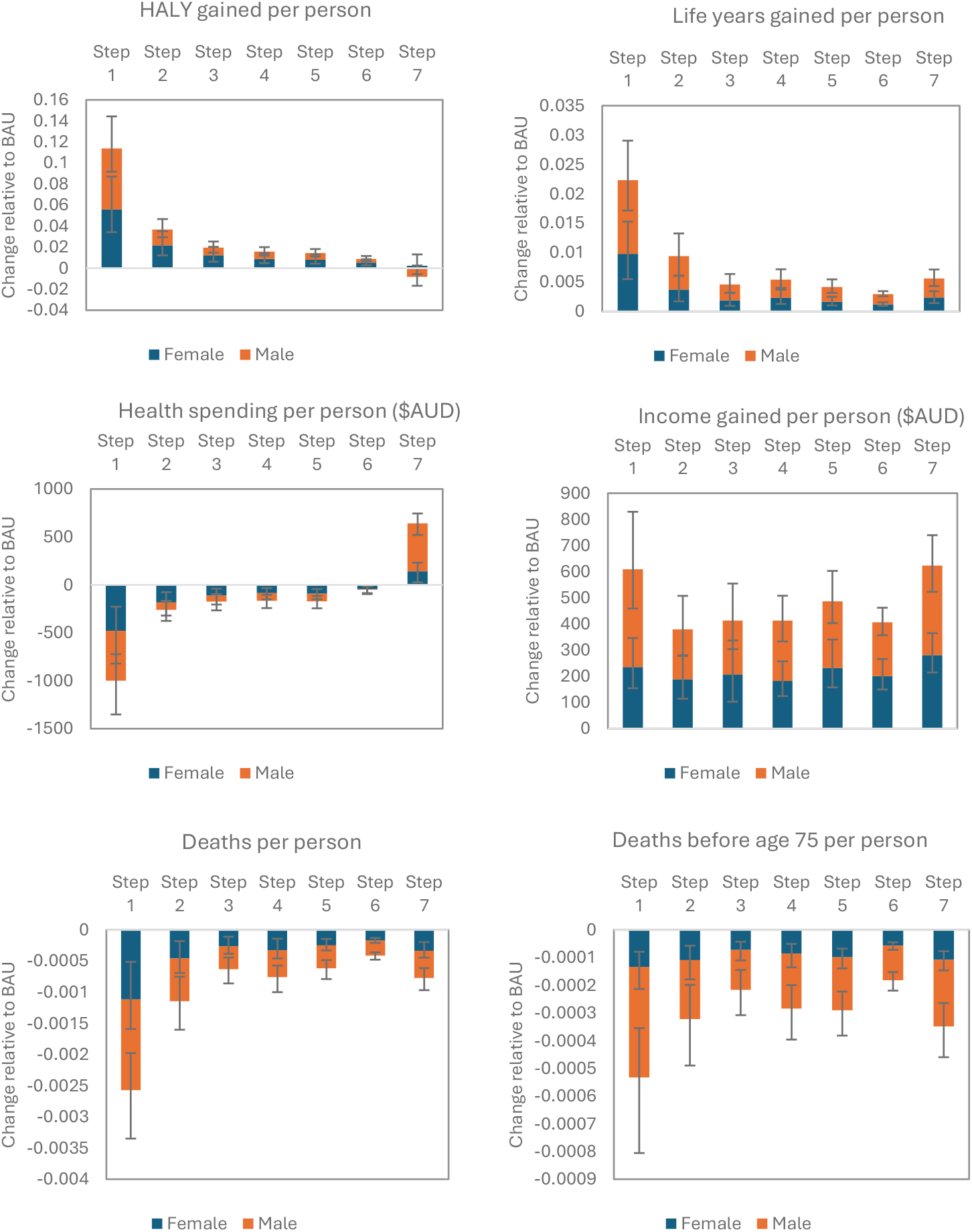
Per person change in HALYS, life years, health spending, income, total deaths, and deaths before age 75, relative to baseline for scenario 5, over 20 years with a 0% discount rate. Note: Scenario involves iteratively moving each physical activity group to the next highest activity level); Per person refers to the number of males and females at baseline in activity group; Groups move independently. Step 1 = 0-600 METs moved to 600-1200, Step 2 = 600-1200 METS to 1200-1800, Step 3 = 1200-1800 METS to 1800-2400, Step 4 = 1800-2400 METS to 2400-3000, Step 5 = 2400-3000 METS to 3000-3600, Step 6 = 3000-3600 METS to 3600-4200, Step 7 = 3600-4200 METS to 4200+.

We now describe key findings by research questions.

### What is the benefit gained from the current level of physical activity undertaken, and how much more health improvements can be made by increasing activity levels?

By modelling the effects of the making everyone inactive (scenario 4), we can see what the benefit gained from the current level of physical activity is (BAU). The current level of physical activity undertaken in Australia contributes to 1.3 million HALYs (679,000 – 2.2 million), defers 28,700 (95% uncertainty interval: 19,100 – 41,100) deaths beyond 2041, and 12,100 (8,180 – 17,900) premature deaths (deaths before the age of 75). As a percent of total deaths expected without any physical activity, this represents a reduction of 0.67% (0.44 – 0.96), and 1.20% (0.81 – 1.77), respectively. Additionally, total income over 20 years would be AUD$16.9 billion (12.0 – 23.2 billion) higher than it would be without doing any activity (0.14%; 0.10 – 0.20), and health spending would be $9.8 billion lower (3.32 – 18.6 billion, or 0.27%; 0.09 – 0.52).

Physical activity levels in Australia increased between 2018 and 2022. By modelling a reversion to the 2018 distribution (scenario 3), we can see what impact this increase in physical activity would have over 20 years, as the BAU comparison reflects the 2022 physical activity level. Overall, the level of activity undertaken in 2022 saves $1.45 billion (655 million to 2.49 billion) in health spending relative to the 2018 activity level and increase HALYs by 151,000 (84,300 – 244,000).

Over 20 years, increasing physical activity further to the theoretical minimum risk level of 4200+ MET-min/week (scenario 1) would increase total HALYS relative to the BAU by 653,000 (230,000 – 1,210,000). As a percent of BAU HALYs, this would be 0.16% (0.06 – 0.30). Total deaths over the 20-year period would decrease by 33,700 (25,700 – 43,700), or -0.78% (-0.60 – -1.02), however deaths before age 75 would decrease by 0.96 % (0.73 – 1.02). All the metrics examined improved by moving the population to the highest level of physical activity, except for total health spending for males. While total health spending for females would decrease by 2.1 billion (-1.01 billion – 5.9 billion), for males it would increase by 1.33 billion (-1.31 billion to 3.51 billion). When physical inactivity is increased for those who are sedentary to meet physical activity guidelines (scenario 2), health spending does reduce for both males and females.

Changes in total deaths, HALYs, income, and health costs per year over 40 years are reported in Figure 1 for each of these physical activity scenarios. While health costs would initially increase in scenario 1 (maximal activity) due to increased injury risk, health costs would decrease relative to the BAU after 8 years and remain lower until 2058. The other scenarios modelled follow the expected patterns, wherein increases in physical activity reduce health spending, and vice versa decreases in activity increase spending. Changes by sex are reported in Supplementary Figure 1. Cumulative changes are reported in Supplementary Figures 3-6.

### What is the negative effect of injuries compared to the positive effect on disease risk?

Disease groups contributing to the overall change in health effects (HALYs) and health spending for males and females are reported in Table 3. The current level of physical activity (i.e. BAU as the scenario compared with everyone physically inactive, scenario 4) has relatively little HALY reduction due to injuries (421 thousand; 344,000 – 507,000) compared to the HALY increase from mental health (1.3 million; 700,000 – 2.1 million), though slightly more than cardiovascular diseases (183,000; 116,000 – 276,000). The current level of physical activity saves the health system 9.8 billion (3.3 – 18.6 billion), with injuries contributing to a 10.2 billion (8.8 – 11.5 billion) increase in spending, and savings from mental health reducing spending by 11.2 billion (5.9 – 19.2 billion). The impact of injuries on health spending for males was 2.6 times that of females.

The impact of injuries due to physical activity is greater in scenario 1(maximum activity) than other scenarios. The increase in health spending for males in scenario 1 was due to the impact of injuries sustained during activity increasing health spending further than the overall reduction in spending on diseases associated with physical inactivity. While the net reduction in health spending over 20 years would be 748 million (-6.9 to 4.5 billion), the effect of injuries due to physical activity in this scenario increase health spending by 16.9 billion (15.1 to 18.6 billion), offset by a 17.3 billion decrease in spending from other diseases. The effect for males is $2.5 billion higher than that of females. The next largest impact on health spending was due to improvements to mental health, which would reduce health spending by $7.96 billion (-4.3 to - 13.5 billion). On the other hand, the contribution of injuries to change in HALYs was less than that of mental health.

The net effect of physical activity on injuries is a combination of both the positive effect of physical activity on bone mineral density and subsequent fractures, and negative effects on injuries during participation. Supplementary Table S6 shows HALYs and spending due to disease groups over 20 years, compared to a sensitivity analysis with no injuries due to physical activity participation (model 2). Model 2 does include injuries prevented through bone mineral density improvements due to physical activity. In this model, spending on injuries (due to low bone mineral density) decreased by $2.3 billion (957 million – 3.9 billion), and HALYs increased by 116,000 (49,800 to 202,000). Therefore, the effect of physical activity on injuries due to participation is greater than is reflected in the injury contribution to total effects, as this reflects the net effect of these pathways.

### At what point does the injury negative effect outweigh the positive disease risk reductions?

By modelling separately each physical activity group shifting to the next highest physical activity group, we can see at which level of physical activity the negative impact of injuries outweighs the beneficial effects on disease risk (scenario 5). For total deaths, deaths before age 75, and years of life gained per person, the net impact of physical activity attenuates as activity level increases but does not reach a point of negative impacts. However, HALY gained per person becomes slightly negative overall in the 4200+ group. For males there is a decrease in HALY in this group, while for females the increase in HALY is negligeable and the lower uncertainty interval crosses the null threshold. The change in health spending per person is neutral in the 3600-4200 MET-min/week group, however spending increases substantially at 4200+ MET-min/week, for both males and females. The effect of injuries on health spending in each group is greater than for females.

### How do these results compare to results obtained from a previously standard list of conditions, or excluding injury risk?

Including injuries caused by physical activity is not common in simulation analyses such as this, and a sensitivity analysis was undertaken to compare the results of this model (all pathways including injuries, model 1), with what would be expected when all of the pathways to disease but no injuries due to activity are included (model 2), and the list of conditions that are more standard in physical activity models (model 3). Table 4 includes a comparison of the main model with alternate models 1 and 2 of impacts due to physical activity each year for 20 years for scenario 1. Over 20 years, the total change in HALYs was highest in model 2 (all disease pathways but no injuries due to activity) and lowest in model 3 (standard conditions). Total deaths were similar between models 1 and 2, through smaller reductions were achieved in model 3. Total income gained was similar between models 1 and 2, with model 3 having less income gains. Health spending reductions were greatest in model 2 and were substantially greater than models 1 and 3. Removing time lags for all conditions (model 4) had relatively minor impacts on most metrics, but improved health spending reductions 4.2 times to $3.1 billion. An additional sensitivity analysis using an alternate method to distribute injuries across physical activity groups, based on total time undertaking activity rather than total MET-min/week, showed no difference to model 1.

## Discussion

This study estimated the heath gains and economic impacts over time in the Australian population under various scenarios of physical activity participation. Our study builds upon previous studies that have modelled the impact of physical activity scenarios (5-7, 21), by also including the effect on physical activity on mental health, through other intermediate risk factors to the diseases associated with them (blood pressure, diabetes and fasting plasma glucose, LDL-cholesterol, and bone mineral density), and onto injuries.

We found that compared to a scenario where everyone was physically inactive, current BAU levels of physical inactivity continued over the next 20 years (without discounting) would gain 1.3 million HALYs; prevent 9.8 billion of health spending, averted 28,700 deaths of any age, and avert 12,100 premature deaths before the age of 75 (1.2% reduction). There is still considerable scope for improvement, and increasing physical activity has substantial impacts on health (HALYs, life years, and avoided death).

An important innovation in our modelling was to include anxiety and depression, and the impacts of physical activity through changing other risk factors. However, the effect of injuries is important as physical activity levels increase, particularly for health expenditure. Compared to the standard conditions linked to physical activity in models, our model expansion increased HALY gains by 54%, decreased health system savings by 90% (because of offsetting impact of injuries), and increased income gains by a third. Increasing the physical activity level of those sedentary to meet physical activity guidelines achieves greater health expenditure improvements than the extreme scenario of everyone at maximal PA level. If injuries were excluded from the model, the estimated benefits for HALYs and health spending increase substantially.

One study using similar modelling techniques for Australia(21) found that over 25 years, if Australia achieved the physical activity guidelines, 227,785 (89,797 – 375,869) HALYs would be gained by reduced anxiety, and 209,199 (108,518 – 313,342) HALYs would be gained by reduced depression. This is higher than the estimated increase in HALYs over 20 years in our model of 168,000 (89,000 to 279,000) for depression and anxiety combined. However, our estimate of reduced health spending for depression and anxiety was similar (at about $1.64 billion (($851 million to $2.86 billion)) over 20 years) than in this study (about $1.8 billion ($691 million to $2.9 billion) over 25 years). Both studies use disease costs derived from the AIHW Disease Expenditure in Australia 2018-19 report(18). The main difference in modelling technique is different activity thresholds for physical activity groups, and the use of an open cohort in our study, and a closed cohort by Wanjau, which may explain some of the differences.

To our knowledge, no other study has modelled both the benefits of physical activity on chronic diseases and the injury risk from participating in physical activity for the whole population. Some studies have included injuries from activity for active transport intervention models(5-7)– in one study it was estimated that while their active transport intervention (replacing short distance car trips with walking, and longer trips with either cycling or public transport) is beneficial for health overall, DALYS from road trauma for pedestrians and cyclists would increase by 7%(6). As a ratio of injury HALYs caused by activity to HALYs averted by increasing activity, this is equivalent to 19% and is similar to the ratios found in our study for the level of activity currently undertaken compared to no activity (25%), though in the scenario where everyone is in the 4200+ MET-min/week group this ratio increases to 54%. This study also modelled exposure to air pollution from increased outside activities (which had very little impact on overall results), which we did not include in our model, as physical activity was specified only by the total amount undertaken, not specifying the type or location.

The injury rate estimated for total injuries was 0.06 (0.04 – 0.09) per 10,000 MET-min/week (or 0.39 per 1,000 MET-hours). This was slightly higher than the injury rate identified in a systematic review of injury rates in children, which found a rate of between 0.15 to 0.27 injuries per 1,000 hours of physical activity(22). Our estimate only relates to adults, who may have a different pattern of injury due to the types of activities undertaken.

## Limitations

The injury risk for physical activity was estimated based on the proportion of total incident injuries that were likely due to physical activity, distributed across the total volume of physical activity undertaken. Estimates of the percent of injuries due to physical activity were based on the percentage of hospitalised injuries by age, sex, and injury type(17), that were identified as being due to physical activity(4). This may not reflect the broader pattern of injuries across Australia, as this represents the more severe cases of injury. However, proportions of injuries identified as being due to activity in hospitals was broadly similar to that of survey data from general practice(23, 24).

The injuries in each physical activity group were modelled as a linear function based on the number of MET-min/week undertaken for that group. However, the relationship of injury with physical activity is more complex than simply a function of exposure. A cohort study of Spanish adults found that while injury rates do increase as time spent active increases, it may not increase linearly(25). There may also be a slightly higher risk of injury for those increasing their level of activity from a lower base, as they are less adapted to doing physical activity, while those who are already at a high level of activity may be well adapted to doing activities, or more aware of the injury risks within the activities they do, and so proactively manage these risks to enable a higher volume of physical activity participation. Injuries may also be less severe among those who have a greater history of participating in physical activity(26, 27). There is insufficient evidence at this point to account for these variations in our model. There may also be variations in injury risk for the same activity undertaken at differing intensities(28, 29) (such as a shorter bout of high intensity physical activity compared to a longer duration of lower intensity). To attempt to evaluate the impact of this bias, we conducted a sensitivity analysis with injury rates that were linear with time undertaken, assuming the average intensity of activity in the 0-600 MET-min/week group was 4 METs, increasing by .5 METs in each group to 7.5 MET in the 4200+ group. Results are similar between these two modelling methods.

The injury risk is not uniform across each activity, and some types of activity have a greater risk of injury than others(4). The contribution of each of these types of physical activity to the total MET-min/week undertaken within each physical activity group may differ, which may bias the results. Further work should be undertaken to determine the risk of physical activity related injury by level and type of activity, so that the model can more appropriately reflect the activity risks within each physical activity group, and the benefits to health can be optimised with lowest injury.

Future models of specific physical activity interventions should be mindful of the context in which the activity is occurring and ensure that injury risks are appropriately reflected in the model. For example, it may be the case that cycling for recreation may have a lower injury risk than cycling for transport(30). This may also assist with identifying areas for injury prevention to improve the trade-off between physical activity benefits and injury risk.

Our study only captured the effect of physical activity increases on the rate of incident disease, and did not include any effect of physical activity on those already diseased (e.g. reduced disease severity, increased remission rates, or reduced case fatality). Secondary and tertiary prevention may be particularly relevant for conditions such as type 2 diabetes, depression, or anxiety. If this was included, the benefits of activity would likely be greater than those estimated in our study. The issue here of including effects of intervention scenarios action through risk factors to impact disease rates beyond just incidence rates is a general one for the whole field of health impact assessment of risk factor interventions – and is an area for future improvement as data permits.

The scenarios modelled assume changes effect the whole population within a physical activity group. However, these results are unlikely to be achieved, as for example some individuals may have low physical activity due to disability, or have very high physical activity due to their occupation.

Underpinning all these results is the assumption that the relative risks are not affected by residual confounding or other issues with exchangeability, and are consistent across age and types of activity within METs levels. Actual relative risks may vary by type of activity undertaken to achieve the risk reductions.

## Conclusion

Increasing physical activity levels in Australia would improve population health even when the risk of injuries due to participation is factored in, reduce health spending, and increase income. The greatest gains are likely to be made by increasing physical activity of those who are sedentary. While benefits do continue to accrue as activity increases, the benefits attenuate and the effect of injuries due to activity begins to reduce the overall benefits of physical activity. More research is needed to determine the risk of injury by level of physical activity, to account for adaptations to each level of activity that might reduce injury risk.

## Data Availability

All data produced in the present study are available upon reasonable request to the authors

## Notes

### Competing Interest Statement

The authors have declared no competing interest.

