## Supplementary material for "Future health gain from increasing physical activity in Australia, including multiple physiological effects of physical activity, and falls and injury risk: A simulation study"

### Calculation of risk factor distributions, disease risks, and disease costs

*Calculation of risk factor distributions, disease risks, and disease costs*

Estimating the dose response of physical activity with other mediating risk factors

For blood pressure, blood glucose, and bone mineral density, the differences in risk factor exposure by level of physical activity were derived from published studies(1). Blood pressure and fasting plasma glucose were modelled using differences in the relative risk of hypertension(2) and diabetes(3), and extrapolating the associated shifts in the risk factor means by level of physical activity.

For LDL-cholesterol, the differences in distribution by physical activity level were calculated using a regression analysis of UK Biobank data and adjusting for all other relevant variables (such as age, diet, or smoking status).

Refer to Supplementary Table S3 for further detail on calculations for each risk factor.

*Estimating business-as-usual risk factor distributions by level of physical activity in Australia*

We attempted to use National Health Survey(4) data to estimate joint distributions of physical activity with (individually) blood pressure, blood glucose, bone mineral density, and cholesterol by sex and age, but data sparsity meant estimates were imprecise and unstable. We instead assumed that risk factor distributions (blood pressure, blood glucose, bone mineral density, and LDL-cholesterol) varied by physical activity group within each sex by age group only by the above estimated causal dose-responses. Note that we assumed the distributions of risk factors by sex and age in BAU did not change into the future (i.e. a static forecast), although as cohorts age, they will change to the next age-groups risk factor(s) distribution(s).

*Incidence, case fatality and remission rates of chronic conditions, and event rates of injuries*

Sex by age rates of all included conditions, diseases and injuries (i.e. those in risk-outcome pairs with the above risk factors) were derived from GBD2021(5) data. Briefly, we used an optimisation procedure to ensure cohort coherence of incidence, case fatality and a combined remission and excess mortality rate (i.e. due to correlated mortality risks) for each chronic disease for 1990 to 2021. The GBD outputs were generated with DISMOD-MR, which uses meta-regression methods and predictors of rates suitable for period analyses to generate DALYs – but is not focused solely on cohort coherence of rates over time. Further, GBD outputs do not include remission rates and excess mortality rates – although initial estimates for the sum of these two rates (that are often quite unstable) can be solved from between year differences in GBD disease prevalence and GBD incidence and cause-specific mortality rates. Hence, the need for extra processing to generate a coherent historic series from 1990 to 2021. Next, we used regression forecasts of these rates into the future for 15 years, then assumed no further change (given increasing uncertainty the further into the future forecasts are made). Details of these methods can be found elsewhere. Reduced version methods were used for acute events (e.g. injury).

The PMSLT required disaggregation of joint distributions of physical activity and mediating risk factors within each sex by age category (above). However, disease rates do not need to be thus disaggregated, as sex by age rates of disease under intervention scenarios are changed using population impact fractions that are percentage changes in disease incidence or event rates given a scenario-generated shift in risk factor distributions (see below).

*Rate ratios of disease incidence and event rates by physical activity (for direct effects) and by other risk factors (for indirect effects)*

We required rate ratios of physical activity to disease and conditions directly for: Anxiety disorders, Bowel cancer, Breast cancer, Coronary heart disease, Dementia, Depressive disorders, Stroke, Type 2 diabetes, and Uterine cancer. Rate ratios were also required for many other diseases and conditions from mediating risk factors, listed in Supplementary Table S3. Relative risks were taken directly from the GBD, when that risk-outcome pair was included in GBD2021(5) or ABDS2024(6). The 2019 GBD(7) relative risks were used due to limited data availability of the GBD 2021 relative risks.

For the novel risk outcome pairs we included (depression and anxiety), we generated dose response relative risk functions ourselves. Relative risks for depression and anxiety were derived from systematic reviews reporting results for ‘active’ compared to ‘not active’(8-10), and were converted to a log-linear dose response using the level of physical activity undertaken in the highest and lowest tertile for each age and sex in Australia. Injury incidence due to physical activity was derived using the fraction of each injury type that is due to physical activity from the AIHW(11, 12), by age and sex, and the total injury incidence for each type of injury from the GBD(5). The total incidence of injuries due to physical activity was converted to a rate per MET-min/week, using total MET-min/week of physical activity undertaken for each age and sex(6). Types of injuries due to physical activity in the model are listed in Supplementary Table S1, while the percentage of each injury type that was due to activity for each age and sex is listed in Supplementary Table S2. The injury rate for total injuries was 0.06 (0.04 – 0.09) per 10,000 MET-min/week.

Some conditions with a direct association from physical activity (coronary heart disease, stroke, and dementia(3, 13)) have meta-analysis generated relative risks that are based on studies that do and do not adjust for mediating risk factors; by erroneously including studies adjusted for mediations between physical activity and disease (e.g. blood pressure), there will be a systematic bias to underestimate the total rate ratio. We corrected for this bias by calculating the percent that each risk factor that was over-adjusted and including this percentage of the risk factor-disease pathway in the total estimate. The percentage used is the average of the percentage of the studies and the percentage of total participations underlying the direct physical activity RR that had adjusted for these mediating risk factors as confounders (Supplementary Table S4).

*Costing data*

The health system expenditure per case of disease by phase (incident year, last year of life from that disease, and otherwise prevalent) was estimated by distributing total expenditure for each disease by age group and sex from AIHW Australian estimates(14) across disease phase (first year of diagnosis, last year of diagnosis if dying of disease, and otherwise prevalent) using both: the incidence, prevalence and case fatality data from the GBD(7); and estimates of relative cost of disease by phase from an analysis of New Zealand data(15). Estimates of income loss due to diseases are unavailable for Australia, so we used NZ estimates(16) rescaled to Australia allowing for differences between Australia and NZ in average per capita income.

***References***

1.	Bourke E, Rawstorn J, Maddison R, Blakely T. The effects of physical inactivity on other risk factors for chronic disease: A systematic review of reviews. Preventive Medicine Reports. 2024;46:102866.

2.	Liu X, Zhang D, Liu Y, Sun X, Han C, Wang B, et al. Dose-Response Association Between Physical Activity and Incident Hypertension: A Systematic Review and Meta-Analysis of Cohort Studies. Hypertension. 2017;69(5):813-20.

3.	Kyu HH, Bachman VF, Alexander LT, Mumford JE, Afshin A, Estep K, et al. Physical activity and risk of breast cancer, colon cancer, diabetes, ischemic heart disease, and ischemic stroke events: systematic review and dose-response meta-analysis for the Global Burden of Disease Study 2013. Bmj. 2016;354:i3857.

4.	Australian Bureau of Statistics. National Health Survey. 2022.

5.	Brauer M, Roth GA, Aravkin AY, Zheng P, Abate KH, Abate YH, et al. Global burden and strength of evidence for 88 risk factors in 204 countries and 811 subnational locations, 1990&#x2013;2021: a systematic analysis for the Global Burden of Disease Study 2021. The Lancet. 2024;403(10440):2162-203.

6.	Australian Institute of Health and Welfare. Australian Burden of Disease Study 2024. Canberra: AIHW; 2024.

7.	Murray CJL, Aravkin AY, Zheng P, Abbafati C, Abbas KM, Abbasi-Kangevari M, et al. Global burden of 87 risk factors in 204 countries and territories, 1990-2019: a systematic analysis for the Global Burden of Disease Study 2019. The Lancet. 2020;396(10258):1223-49.

8.	Wanjau MN, Möller H, Haigh F, Milat A, Hayek R, Lucas P, et al. Physical Activity and Depression and Anxiety Disorders: A Systematic Review of Reviews and Assessment of Causality. AJPM Focus. 2023;2(2):100074.

9.	Schuch FB, Stubbs B, Meyer J, Heissel A, Zech P, Vancampfort D, et al. Physical activity protects from incident anxiety: A meta-analysis of prospective cohort studies. Depress Anxiety. 2019;36(9):846-58.

10.	Schuch FB, Vancampfort D, Firth J, Rosenbaum S, Ward PB, Silva ES, et al. Physical Activity and Incident Depression: A Meta-Analysis of Prospective Cohort Studies. Am J Psychiatry. 2018;175(7):631-48.

11.	Australian Institute of Health and Welfare. Sports injury in Australia. Canberra: AIHW; 2024.

12.	Australian Institute of Health and Welfare. Injury in Australia. Canberra: AIHW; 2024.

13.	Blondell SJ, Hammersley-Mather R, Veerman JL. Does physical activity prevent cognitive decline and dementia?: A systematic review and meta-analysis of longitudinal studies. BMC Public Health. 2014;14:510.

14.	Australian Institute of Health and Welfare. Disease expenditure in Australia 2018-19. Canberra: AIHW; 2021.

15.	Blakely T, Kvizhinadze G, Atkinson J, Dieleman J, Clarke P. Health system costs for individual and comorbid noncommunicable diseases: An analysis of publicly funded health events from New Zealand. PLoS Med. 2019;16(1):e1002716.

16.	Blakely T, Sigglekow F, Irfan M, Mizdrak A, Dieleman J, Bablani L, et al. Disease-related income and economic productivity loss in New Zealand: A longitudinal analysis of linked individual-level data. PLoS Med. 2021;18(11):e1003848.

### Supplementary tables and figures

**Supplementary Table S1: Conditions and injuries either prevented or caused by physical activity.**

| Physical activity pathway | Condition or injury |
| --- | --- |
| Preventive | Anxiety disorders, Bowel cancer, Breast cancer, Coronary heart disease, Dementia, Depressive disorders, Stroke, Type 2 diabetes, Uterine cancer |
| Causative | Dislocation of hip, Dislocation of knee, Dislocation of shoulder, Drowning and nonfatal submersion, Fracture of hip, Fracture of clavicle, scapula, or humerus, Asphyxiation, Severe chest Injury, Crush injury, Internal hemorrhage in abdomen and pelvis, Fracture of face bones, Fracture of femur, other than femoral neck, Fracture of foot bones except ankle, Fracture of hand (wrist and other distal part of hand), Fracture of pelvis, Fracture of radius and/or ulna, Fracture of skull, Fracture of sternum and/or fracture of one or more ribs, Fracture of vertebral column, Injury to eyes, Nerve injury, Multiple fractures, dislocations, crashes, wounds, pains, and strains, Superficial injury of any part of the body, Contusion in any part of the body, Muscle and tendon injuries, including sprains and strains lesser dislocations, Open wound(s), Spinal cord lesion below neck level, Spinal cord lesion at neck level, Fracture of patella, tibia or fibula, or ankle, Minor TBI, Moderate/Severe TBI |

Supplementary Table S2: Percent of each injury type due to physical activity, by sex and age

|  |  | Age group | | | |
| --- | --- | --- | --- | --- | --- |
| **Sex** | **Injury type** | **20–24** | **25–44** | **45–64** | **65+** |
| Males | Fracture | 35% | 25% | 13% | 4% |
|  | Dislocation | 34% | 26% | 18% | 7% |
|  | Soft-tissue injury | 27% | 15% | 10% | 9% |
|  | Open wound | 13% | 7% | 4% | 1% |
|  | Intracranial injury | 31% | 27% | 13% | 2% |
|  | Nerve injury | 8% | 5% | 3% | 3% |
|  | Burn | 5% | 3% | 1% | 1% |
|  | Superficial injury | 21% | 13% | 6% | 1% |
|  | Poisoning or toxic effect | 1% | 0% | 0% | 0% |
|  | Amputation | 5% | 2% | 1% | 1% |
|  | Crushing injury | 13% | 8% | 4% | 5% |
|  | Internal organs | 30% | 23% | 11% | 5% |
|  | Blood vessels | 5% | 2% | 2% | 1% |
|  | Foreign object (through orifice) | 1% | 0% | 0% | 0% |
|  | Other specified and/or multiple injuries | 29% | 17% | 10% | 2% |
| Females | Fracture | 40% | 22% | 8% | 2% |
|  | Dislocation | 25% | 15% | 10% | 3% |
|  | Soft-tissue injury | 19% | 14% | 11% | 9% |
|  | Open wound | 7% | 4% | 3% | 1% |
|  | Intracranial injury | 21% | 19% | 12% | 2% |
|  | Nerve injury | 7% | 4% | 4% | 5% |
|  | Burn | 3% | 2% | 2% | 1% |
|  | Superficial injury | 9% | 6% | 4% | 1% |
|  | Poisoning or toxic effect | 0% | 0% | 0% | 0% |
|  | Amputation | 4% | 2% | 1% | 1% |
|  | Crushing injury | 6% | 4% | 2% | 5% |
|  | Internal organs | 14% | 10% | 8% | 4% |
|  | Blood vessels | 2% | 2% | 1% | 1% |
|  | Foreign object (through orifice) | 0% | 0% | 0% | 0% |
|  | Other specified and/or multiple injuries | 14% | 9% | 8% | 2% |

Supplementary Table S3: Effect measures used to estimate difference in intermediate risk factors by level of physical activity, and conditions associated with each risk factor

Note: Table is adapted from “The total burden of physical inactivity in a comparative risk assessment: including indirect effects on chronic disease, and direct effects on mental health” by Bourke, E, Maddison, R, and Blakely, T, 2025. Conditions in bold are also linked directly to physical inactivity and have a direct RR component.

| Intermediate risk factor | Source for physical activity effect on risk factor | Modelling method to calculate difference in risk factor mean by activity level | Conditions linked to each risk factor (conditions in bold are those included in standard models) | Condition relative risk data sources |
| --- | --- | --- | --- | --- |
| **High blood pressure** | Liu et al, 2017, RR 0.94 per 600 MET-min/week reduction in hypertension | Population average difference in hypertension by physical activity level calculated from hypertension RR, and prevalence of hypertension (based on BP >140mmHg) using the mean and standard deviation (SD) of BP distribution, and assumed normal distribution. Inverse normal distribution used to calculate the shift in mean BP (holding SD constant) required to achieve the required percentage change hypertensive for each physical activity level. Average difference in mean by activity group applied to age and sex specific means. | Aortic aneurysm, atrial fibrillation and flutter, cardiomyopathy, chronic kidney disease, **coronary heart disease**, **dementia**, hypertensive heart disease, inflammatory heart disease, other cardiovascular diseases, peripheral vascular disease, rheumatic heart disease, **stroke**, non-rheumatic valvular disease | GBD 2019 (from APSC Collaboration 2003, Prospective Studies Collaboration 2002, and Rapsomaniki et al, 2014), and Mulligan et al, 2023 (for dementia) |
| **High fasting plasma glucose** | Kyu et al, 2016, diabetes dose response | Population average difference in diabetes by physical activity level calculated from diabetes RR, and prevalence of diabetes (based on FPG >7.0 mmol/L) using the mean and standard deviation of FPG distribution, and assumed normal distribution. Inverse normal distribution used to calculate the shift in mean fasting plasma glucose (holding SD constant) required to achieve the required percentage change in diabetic for each physical activity level. Average difference in mean by activity group applied to age and sex specific means. | Chronic kidney disease, **coronary heart disease**, **stroke** | GBD 2019 (from Singh et al, 2013) |
| **High LDL-cholesterol** | Analysis of UK-Biobank data. Reduction of 0.017 (0.013 – 0.022) mmol/L per log unit MET-min/week | Decrease in mean LDL-cholesterol per ln(MET-mins/week) in each activity group applied to age and sex specific means. The regression model used to derive log-linear dose response included age, sex, BMI, education status, dietary score, and smoking status as covariates. | **Coronary heart disease, stroke** | GBD 2019 (from Singh et al, 2013) |
| **Low bone mineral density** | Pinheiro et al, 2020. Increase of 0.15 (0.05 – 0.25) standard deviations | We assumed the MET-min/week difference in the ‘inactive’ and ‘active’ intervention groups reflected the difference in METs between those undertaking 0-600 MET-min/week and anything above 600 in National Health Survey 2022 data. A log linear dose response was calculated, with the increase in mean BMD for each physical activity group calculated based on MET-min/week undertaken, assuming that bone mineral density increases have lower marginal gains at higher activity levels. | Hip fracture, humerus fracture, other fractures, tibia and ankle fracture | GBD 2019 (from Johnell et al, 2005) |
| **Type 2 diabetes** | Kyu et al, 2016, diabetes dose response | Type 2 diabetes prevalence is part of the exposure to high fasting plasma glucose, and has further conditions associated with it. We therefore model type 2 diabetes as a separate risk factor. Difference in prevalence of diabetes by physical activity level calculated from RR of diabetes for each physical activity level, directly applied to age and sex diabetes prevalence. | Bladder cancer, **bowel cancer**, **breast cancer**, cataract and other lens disorders, chronic kidney disease, **dementia**, glaucoma, liver cancer, lung cancer, ovarian cancer, pancreatic cancer, peripheral vascular disease | GBD 2019 (from Singh et al, 2013) |

Supplementary Table S4: Summary of mediator adjustments in studies underlying relative risk estimates from meta-analyses

Note: Table is adapted from “The total burden of physical inactivity in a comparative risk assessment: including indirect effects on chronic disease, and direct effects on mental health” by Bourke, E, Maddison, R, and Blakely, T, 2025.

| Systematic review | N (studies) | N (participants) | Percent (studies) | Percent (participants) | Midpoint used to model pathway |
| --- | --- | --- | --- | --- | --- |
| **Coronary heart disease, Kyu et al, 2016** |  |  |  |  |  |
| *Total studies in meta-analysis* | *43* | *1,838,930* | *100%* | *100%* | *100%* |
| No mediator adjusted for | 20 | 1,393,016 | 47% | 76% | 61% |
| Any mediator adjusted for | 23 | 445,914 | 53% | 24% | 39% |
| Blood pressure | 20 | 338,891 | 47% | 18% | 32% |
| Cholesterol | 17 | 317,602 | 40% | 17% | 28% |
| Fasting plasma glucose | 1 | 4,889 | 2% | 0% | 1% |
| Type 2 diabetes | 14 | 332,510 | 33% | 18% | 25% |
| **Stroke, Kyu et al, 2016** |  |  |  |  |  |
| *Total studies in meta-analysis* | *26* | *1,573,231* | *100%* | *100%* | *100%* |
| No mediator adjusted for | 12 | 1,236,505 | 46% | 79% | 62% |
| Any mediator adjusted for | 14 | 336,726 | 54% | 21% | 38% |
| Blood pressure | 13 | 329,285 | 50% | 21% | 35% |
| Cholesterol | 8 | 296,413 | 31% | 19% | 25% |
| Fasting plasma glucose | 2 | 93,552 | 8% | 6% | 7% |
| Type 2 diabetes | 8 | 219,603 | 31% | 14% | 22% |
| **Dementia, Blondell et al, 2014** |  |  |  |  |  |
| *Total studies in meta-analysis* | *39* | *87,374* | *100%* | *100%* | *100%* |
| No mediator adjusted for | 22 | 32,161 | 56% | 37% | 47% |
| Any mediator adjusted for | 17 | 55,213 | 44% | 63% | 53% |
| Blood pressure | 14 | 42,466 | 36% | 49% | 42% |
| Cholesterol | 10 | 33,165 | 26% | 38% | 32% |
| Type 2 diabetes | 14 | 39,132 | 36% | 45% | 40% |

**Supplementary Table S5: Time lags implemented for each condition.**

| **Condition** | **Time Lag** |
| --- | --- |
| "anxiety_disorder" | Nil |
| "aortic_aneurysm" | Short (5 years) |
| "asphyxiation" | Nil |
| "atrial_fib_and_flutter" | Short (5 years) |
| "bladder_cancer" | Long (20 years) |
| "blindness_vision_loss" | Short (5 years) |
| "breast_cancer" | Long (20 years) |
| "cardiomyopathy" | Short (5 years) |
| "cardiomyopathy_acute" | Short (5 years) |
| "chest_injury" | Nil |
| "chronic_kidney_disease" | Long (20 years) |
| "colorectal_cancer" | Long (20 years) |
| "contusions" | Nil |
| "crush_injury" | Nil |
| "dementia" | Long (20 years) |
| "depressive_disorder" | Nil |
| "diabetes_mellitus" | Short (5 years) |
| "dislocations_hip" | Nil |
| "dislocations_knee" | Nil |
| "dislocations_shoulder" | Nil |
| "drowning_and_submersion" | Nil |
| "fracture_hip" | Nil |
| "fracture_humerus" | Nil |
| "fracture_tibia_and_ankle" | Nil |
| "fractures_face" | Nil |
| "fractures_femur" | Nil |
| "fractures_foot" | Nil |
| "fractures_hand" | Nil |
| "fractures_pelvis" | Nil |
| "fractures_radius" | Nil |
| "fractures_skull" | Nil |
| "fractures_sternum" | Nil |
| "fractures_verbetral_col" | Nil |
| "hypertensive_heart_disease" | Long (20 years) |
| "ihd" | Short (5 years) |
| "injury_eye" | Nil |
| "injury_nerve" | Nil |
| "internal_hemorrhage" | Nil |
| "intracerebral_hemorrhage" | Short (5 years) |
| "liver_cancer" | Long (20 years) |
| "lung_cancer" | Long (20 years) |
| "muscle_and_tendon" | Nil |
| "nr_calcific_aortic_valve_disease" | Short (5 years) |
| "open_wounds" | Nil |
| "other_injuries_multiple" | Nil |
| "other_injuries_superficial" | Nil |
| "ovarian_cancer" | Long (20 years) |
| "pancreatic_cancer" | Long (20 years) |
| "peripheral_artery_disease" | Long (20 years) |
| "rheumatic_heart_disease" | Long (20 years) |
| "sci_below_neck" | Nil |
| "sci_neck" | Nil |
| "tbi_minor" | Nil |
| "tbi_moderate_sev" | Nil |
| "uterine_cancer" | Long (20 years) |

**Supplementary Figure S1: Change in health spending for males and females relative to BAU for 4 main intervention scenarios, each year for 40 years with 0% discount rate**

Note: 1) Maximal activity: 100% of each age and sex reached the highest physical activity category (4200+ MET-min/week), 2) Meet guidelines: everyone in the 0-600 MET-min/week group moved to the 600-1200 MET-min/week group (meeting minimum total physical activity guidelines), 3) Revert to 2018: the population reverting to the lesser 2018 physical activity level for each age and sex, 4) Everyone inactive: all sex by age groups set to 0-600 MET-min/week

Change in health spending, Females

Change in health spending (millions)

|  | 1. Maximal activity | 2. Meet guidelines | 3. Revert to 2018 | 4. Everyone inactive |  |
| --- | --- | --- | --- | --- | --- |
| 2021 | 101.33 | -40.9394 | 11.5886 | 142.903 | 0 |
| 2022 | 80.4835 | -43.753 | 14.7235 | 159.253 | 0 |
| 2023 | 56.2207 | -46.1733 | 18.4491 | 178.398 | 0 |
| 2024 | 28.2072 | -48.1524 | 22.6005 | 198.511 | 0 |
| 2025 | 0.895608 | -49.584 | 26.579 | 219.112 | 0 |
| 2026 | -25.1968 | -50.6577 | 30.3581 | 237.322 | 0 |
| 2027 | -48.2533 | -51.4507 | 33.888 | 254.113 | 0 |
| 2028 | -70.1882 | -51.8837 | 37.0788 | 270.235 | 0 |
| 2029 | -91.2943 | -52.436 | 40.081 | 284.87 | 0 |
| 2030 | -110.552 | -52.7619 | 43.0254 | 299.063 | 0 |
| 2031 | -129.59 | -52.9803 | 45.6877 | 313.239 | 0 |
| 2032 | -147.092 | -53.1696 | 48.0932 | 327.298 | 0 |
| 2033 | -164.33 | -53.4615 | 50.4341 | 340.276 | 0 |
| 2034 | -180.668 | -53.6103 | 52.5995 | 351.702 | 0 |
| 2035 | -195.45 | -53.762 | 54.533 | 362.78 | 0 |
| 2036 | -210.569 | -53.9507 | 56.2885 | 373.435 | 0 |
| 2037 | -221.997 | -53.7704 | 57.7018 | 381.428 | 0 |
| 2038 | -232.898 | -53.253 | 58.814 | 389.006 | 0 |
| 2039 | -241.272 | -52.808 | 59.5244 | 395.309 | 0 |
| 2040 | -247.086 | -52.1686 | 59.8574 | 399.003 | 0 |
| 2041 | -251.862 | -50.6711 | 59.8223 | 398.113 | 0 |
| 2042 | -256.387 | -49.1139 | 59.8947 | 397.194 | 0 |
| 2043 | -260.823 | -47.5282 | 59.9024 | 396.453 | 0 |
| 2044 | -263.715 | -46.1457 | 59.6624 | 394.639 | 0 |
| 2045 | -264.451 | -44.6742 | 59.1613 | 391.127 | 0 |
| 2046 | -261.154 | -42.9098 | 57.8664 | 384.229 | 0 |
| 2047 | -257.594 | -41.2091 | 56.6101 | 377.319 | 0 |
| 2048 | -252.676 | -39.5132 | 55.1919 | 369.339 | 0 |
| 2049 | -248.524 | -37.7095 | 53.7906 | 360.761 | 0 |
| 2050 | -242.513 | -36.21 | 52.2967 | 352.455 | 0 |
| 2051 | -235.835 | -34.8216 | 50.7291 | 343.381 | 0 |
| 2052 | -227.575 | -33.2489 | 48.9699 | 333.795 | 0 |
| 2053 | -219.101 | -31.7428 | 47.4342 | 324.234 | 0 |
| 2054 | -209.982 | -30.3242 | 45.8958 | 314.509 | 0 |
| 2055 | -200.15 | -28.8883 | 44.3322 | 303.624 | 0 |
| 2056 | -190.855 | -27.7037 | 42.6256 | 292.992 | 0 |
| 2057 | -180.599 | -26.394 | 40.8528 | 281.477 | 0 |
| 2058 | -170.329 | -25.0377 | 39.1123 | 270.419 | 0 |
| 2059 | -160.51 | -23.8675 | 37.3195 | 259.372 | 0 |
| 2060 | -149.234 | -22.67 | 35.4187 | 248.254 | 0 |

Change in health spending, Males

Change in health spending (millions)

|  | 1. Maximal activity | 2. Meet guidelines | 3. Revert to 2018 | 4. Everyone inactive |  |
| --- | --- | --- | --- | --- | --- |
| 2021 | 315.309 | -26.8786 | -8.80715 | -98.6598 | 0 |
| 2022 | 282.865 | -31.3988 | -3.71222 | -58.9047 | 0 |
| 2023 | 248.668 | -35.2576 | 1.90445 | -15.9195 | 0 |
| 2024 | 213.017 | -38.5129 | 7.71811 | 27.418 | 0 |
| 2025 | 180.457 | -40.7283 | 13.086 | 65.1114 | 0 |
| 2026 | 150.576 | -42.366 | 18.0401 | 100.039 | 0 |
| 2027 | 123.264 | -43.6051 | 22.6434 | 132.452 | 0 |
| 2028 | 98.8538 | -44.4302 | 26.9671 | 161.642 | 0 |
| 2029 | 75.5301 | -44.9786 | 30.8631 | 188.828 | 0 |
| 2030 | 53.974 | -45.3452 | 34.618 | 214.236 | 0 |
| 2031 | 33.0074 | -45.4805 | 38.046 | 238.707 | 0 |
| 2032 | 13.4472 | -45.4587 | 41.2243 | 260.076 | 0 |
| 2033 | -4.28421 | -45.3352 | 44.0375 | 280.041 | 0 |
| 2034 | -22.0871 | -45.3609 | 46.5783 | 300.224 | 0 |
| 2035 | -39.0871 | -45.4141 | 48.7613 | 320.363 | 0 |
| 2036 | -54.2164 | -45.4342 | 50.585 | 338.249 | 0 |
| 2037 | -67.1254 | -45.1666 | 51.9471 | 354.39 | 0 |
| 2038 | -78.7472 | -44.6301 | 52.6675 | 366.68 | 0 |
| 2039 | -89.2395 | -44.0401 | 53.1649 | 377.719 | 0 |
| 2040 | -97.9247 | -43.3708 | 53.2175 | 384.788 | 0 |
| 2041 | -112.581 | -42.7252 | 53.5301 | 397.314 | 0 |
| 2042 | -128.566 | -42.0477 | 53.9512 | 410.377 | 0 |
| 2043 | -145.112 | -41.289 | 54.4581 | 424.202 | 0 |
| 2044 | -159.017 | -40.3781 | 54.6685 | 436.116 | 0 |
| 2045 | -170.928 | -39.2166 | 54.7159 | 444.679 | 0 |
| 2046 | -172.524 | -37.994 | 54.0268 | 443.224 | 0 |
| 2047 | -173.432 | -36.5641 | 53.1605 | 441.084 | 0 |
| 2048 | -173.49 | -35.0104 | 52.1656 | 438.069 | 0 |
| 2049 | -173.53 | -33.4458 | 51.1191 | 432.904 | 0 |
| 2050 | -172.344 | -31.7625 | 50.071 | 426.467 | 0 |
| 2051 | -170.677 | -30.1962 | 48.5626 | 418.556 | 0 |
| 2052 | -168.959 | -28.5028 | 47.2357 | 410.532 | 0 |
| 2053 | -165.992 | -26.6691 | 45.9292 | 401.993 | 0 |
| 2054 | -162.704 | -25.042 | 44.6127 | 391.188 | 0 |
| 2055 | -158.51 | -23.1362 | 43.1912 | 378.627 | 0 |
| 2056 | -150.839 | -21.5964 | 42.1994 | 366.113 | 0 |
| 2057 | -142.466 | -19.9473 | 41.1219 | 352.006 | 0 |
| 2058 | -134.26 | -18.3129 | 40.1626 | 337.466 | 0 |
| 2059 | -125.441 | -16.7665 | 38.9864 | 323.545 | 0 |
| 2060 | -116.582 | -15.2555 | 37.8784 | 306.786 | 0 |

**Supplementary Figure S2: Cumulative change in HALY for both sexes.**

Cumulative difference in HALY

Change from BAU (thousands)

|  | 1. Maximal activity | 2. Meet guidelines | 3. Revert to 2018 | 4. Everyone inactive |  |
| --- | --- | --- | --- | --- | --- |
| 2021 | 5.3838 | 7.55726 | -3.99568 | -42.1553 | 0 |
| 2022 | 11.6878 | 15.2485 | -8.13279 | -85.1715 | 0 |
| 2023 | 19.5052 | 23.1366 | -12.4843 | -129.48 | 0 |
| 2024 | 29.4574 | 31.285 | -17.1322 | -175.567 | 0 |
| 2025 | 41.9681 | 39.7438 | -22.1098 | -223.795 | 0 |
| 2026 | 57.3591 | 48.5537 | -27.4718 | -274.461 | 0 |
| 2027 | 75.8293 | 57.7397 | -33.2339 | -327.784 | 0 |
| 2028 | 97.5501 | 67.317 | -39.4262 | -383.836 | 0 |
| 2029 | 122.731 | 77.2878 | -46.0716 | -442.692 | 0 |
| 2030 | 151.553 | 87.6664 | -53.1765 | -504.532 | 0 |
| 2031 | 184.066 | 98.4495 | -60.7475 | -569.354 | 0 |
| 2032 | 220.393 | 109.635 | -68.8022 | -637.272 | 0 |
| 2033 | 260.607 | 121.229 | -77.343 | -708.439 | 0 |
| 2034 | 304.756 | 133.202 | -86.3734 | -782.635 | 0 |
| 2035 | 352.941 | 145.567 | -95.9205 | -859.93 | 0 |
| 2036 | 405.155 | 158.301 | -105.978 | -940.379 | 0 |
| 2037 | 461.185 | 171.379 | -116.511 | -1023.75 | 0 |
| 2038 | 521.076 | 184.774 | -127.563 | -1110.19 | 0 |
| 2039 | 584.846 | 198.468 | -139.122 | -1199.61 | 0 |
| 2040 | 652.432 | 212.457 | -151.171 | -1291.99 | 0 |
| 2041 | 723.495 | 226.621 | -163.644 | -1386.49 | 0 |
| 2042 | 797.977 | 240.948 | -176.511 | -1482.96 | 0 |
| 2043 | 875.781 | 255.436 | -189.766 | -1581.34 | 0 |
| 2044 | 956.956 | 270.044 | -203.408 | -1681.59 | 0 |
| 2045 | 1041.26 | 284.787 | -217.424 | -1783.81 | 0 |
| 2046 | 1128.53 | 299.614 | -231.807 | -1887.58 | 0 |
| 2047 | 1218.66 | 314.518 | -246.558 | -1992.89 | 0 |
| 2048 | 1311.55 | 329.508 | -261.645 | -2099.67 | 0 |
| 2049 | 1407.2 | 344.572 | -277.07 | -2208.02 | 0 |
| 2050 | 1505.48 | 359.697 | -292.829 | -2317.79 | 0 |
| 2051 | 1606.46 | 374.915 | -308.866 | -2428.91 | 0 |
| 2052 | 1709.96 | 390.213 | -325.152 | -2541.35 | 0 |
| 2053 | 1815.9 | 405.566 | -341.678 | -2654.88 | 0 |
| 2054 | 1924.22 | 420.973 | -358.423 | -2769.7 | 0 |
| 2055 | 2034.68 | 436.412 | -375.388 | -2885.73 | 0 |
| 2056 | 2147.14 | 451.873 | -392.525 | -3002.73 | 0 |
| 2057 | 2261.48 | 467.341 | -409.813 | -3120.57 | 0 |
| 2058 | 2377.58 | 482.826 | -427.218 | -3239.21 | 0 |
| 2059 | 2495.41 | 498.319 | -444.747 | -3358.54 | 0 |
| 2060 | 2614.83 | 513.823 | -462.392 | -3478.37 | 0 |

**Supplementary Figure S3: Cumulative change in health spending for both sexes.**

Cumulative difference in health spending

Change from BAU (millions)

|  | 1. Maximal activity | 2. Meet guidelines | 3. Revert to 2018 | 4. Everyone inactive |  |
| --- | --- | --- | --- | --- | --- |
| 2021 | 416.275 | -68.077 | 2.76438 | 43.5011 | 0 |
| 2022 | 779.367 | -143.233 | 13.6614 | 144.191 | 0 |
| 2023 | 1084.55 | -224.749 | 34.0404 | 306.618 | 0 |
| 2024 | 1325.84 | -311.422 | 64.4191 | 532.143 | 0 |
| 2025 | 1504.69 | -401.874 | 104.425 | 816.629 | 0 |
| 2026 | 1628.45 | -495.08 | 153.156 | 1153.63 | 0 |
| 2027 | 1702.96 | -590.434 | 210.019 | 1539.9 | 0 |
| 2028 | 1730.1 | -686.856 | 274.57 | 1971.13 | 0 |
| 2029 | 1713.55 | -784.021 | 346.008 | 2445.28 | 0 |
| 2030 | 1654.85 | -881.745 | 423.784 | 2957.98 | 0 |
| 2031 | 1557.66 | -979.838 | 507.532 | 3509.51 | 0 |
| 2032 | 1423.12 | -1078.55 | 596.84 | 4096.92 | 0 |
| 2033 | 1252.3 | -1177.59 | 691.227 | 4719.04 | 0 |
| 2034 | 1047.69 | -1276.86 | 790.355 | 5370.29 | 0 |
| 2035 | 811.397 | -1376.38 | 893.821 | 6052.55 | 0 |
| 2036 | 546.059 | -1476.04 | 1000.82 | 6763.95 | 0 |
| 2037 | 255.856 | -1575.2 | 1110.7 | 7499.89 | 0 |
| 2038 | -57.1472 | -1673.59 | 1222.48 | 8255.3 | 0 |
| 2039 | -386.533 | -1770.71 | 1335.53 | 9026.92 | 0 |
| 2040 | -731.595 | -1866.7 | 1448.87 | 9809.22 | 0 |
| 2041 | -1094.56 | -1960.59 | 1562.66 | 10603.8 | 0 |
| 2042 | -1477 | -2052.29 | 1676.59 | 11411.2 | 0 |
| 2043 | -1880.44 | -2141.93 | 1790.86 | 12232.8 | 0 |
| 2044 | -2301.86 | -2229.06 | 1905.11 | 13063.2 | 0 |
| 2045 | -2733.79 | -2313.59 | 2018.94 | 13900.1 | 0 |
| 2046 | -3165.44 | -2394.92 | 2130.82 | 14729.2 | 0 |
| 2047 | -3594.93 | -2473.01 | 2240.21 | 15548.2 | 0 |
| 2048 | -4021.33 | -2547.64 | 2347 | 16356 | 0 |
| 2049 | -4440.85 | -2618.92 | 2451.3 | 17150.2 | 0 |
| 2050 | -4853.96 | -2686.85 | 2553.04 | 17929.6 | 0 |
| 2051 | -5259.48 | -2751.14 | 2651.89 | 18691.8 | 0 |
| 2052 | -5655.85 | -2812.37 | 2747.77 | 19434.5 | 0 |
| 2053 | -6041.72 | -2870.38 | 2840.7 | 20155.5 | 0 |
| 2054 | -6415.09 | -2925.41 | 2930.92 | 20854.8 | 0 |
| 2055 | -6774.67 | -2977.55 | 3018.36 | 21532.1 | 0 |
| 2056 | -7117.59 | -3026.9 | 3103.17 | 22186.9 | 0 |
| 2057 | -7441.59 | -3073.34 | 3185.28 | 22817.2 | 0 |
| 2058 | -7746.55 | -3116.92 | 3264.41 | 23421 | 0 |
| 2059 | -8033.47 | -3157.74 | 3340.71 | 23999.7 | 0 |
| 2060 | -8301.37 | -3195.59 | 3414.16 | 24552.8 | 0 |

**Supplementary Figure S4: Cumulative change in income for both sexes.**

Cumulative difference in Income

Change from BAU (millions)

|  | 1. Maximal activity | 2. Meet guidelines | 3. Revert to 2018 | 4. Everyone inactive |  |
| --- | --- | --- | --- | --- | --- |
| 2021 | 62.2903 | 13.9163 | -11.1766 | -106.238 | 0 |
| 2022 | 179.857 | 31.2885 | -30.1681 | -267.168 | 0 |
| 2023 | 372.481 | 53.3991 | -59.1096 | -501.769 | 0 |
| 2024 | 653.188 | 80.7389 | -99.1099 | -820.248 | 0 |
| 2025 | 1027.49 | 113.436 | -149.893 | -1229.7 | 0 |
| 2026 | 1497.11 | 151.549 | -210.997 | -1730.97 | 0 |
| 2027 | 2061.67 | 194.834 | -282.289 | -2322.18 | 0 |
| 2028 | 2719.31 | 243.022 | -363.693 | -3001.75 | 0 |
| 2029 | 3468.87 | 295.874 | -455.216 | -3769.4 | 0 |
| 2030 | 4311.55 | 353.356 | -557.37 | -4624.4 | 0 |
| 2031 | 5244.21 | 415.052 | -670.032 | -5565.67 | 0 |
| 2032 | 6260.85 | 480.556 | -792.094 | -6587.45 | 0 |
| 2033 | 7354.18 | 549.431 | -922.013 | -7680.84 | 0 |
| 2034 | 8520.6 | 621.503 | -1058.68 | -8840.11 | 0 |
| 2035 | 9748.7 | 696.063 | -1199.84 | -10052.5 | 0 |
| 2036 | 11036.5 | 772.895 | -1344.71 | -11315 | 0 |
| 2037 | 12385.3 | 852.147 | -1494 | -12631.1 | 0 |
| 2038 | 13796.2 | 933.769 | -1647.89 | -14002.5 | 0 |
| 2039 | 15270.4 | 1018.05 | -1806.8 | -15429 | 0 |
| 2040 | 16812.5 | 1105.08 | -1971.26 | -16915.2 | 0 |
| 2041 | 18420 | 1194.67 | -2141.4 | -18458.4 | 0 |
| 2042 | 20092.5 | 1286.83 | -2317.25 | -20057.4 | 0 |
| 2043 | 21823.4 | 1381.26 | -2498.67 | -21706.8 | 0 |
| 2044 | 23608.3 | 1477.67 | -2685.38 | -23402.5 | 0 |
| 2045 | 25434.3 | 1575.63 | -2876.18 | -25133.1 | 0 |
| 2046 | 27289.7 | 1673.86 | -3070.19 | -26886.1 | 0 |
| 2047 | 29164.8 | 1772.01 | -3266.15 | -28653.2 | 0 |
| 2048 | 31056.1 | 1870.03 | -3463.52 | -30432.4 | 0 |
| 2049 | 32956.3 | 1967.61 | -3661.49 | -32217.9 | 0 |
| 2050 | 34859.6 | 2064.53 | -3859.47 | -34005.5 | 0 |
| 2051 | 36765.4 | 2160.88 | -4057.13 | -35792.3 | 0 |
| 2052 | 38666.2 | 2256.6 | -4253.91 | -37572.8 | 0 |
| 2053 | 40555 | 2351.33 | -4449.04 | -39341.1 | 0 |
| 2054 | 42423.2 | 2444.7 | -4641.65 | -41091.4 | 0 |
| 2055 | 44264.1 | 2536.48 | -4831.03 | -42818.1 | 0 |
| 2056 | 46078.5 | 2626.84 | -5017.34 | -44520.5 | 0 |
| 2057 | 47859.9 | 2715.47 | -5200.07 | -46196 | 0 |
| 2058 | 49606.9 | 2802.27 | -5379.08 | -47842.2 | 0 |
| 2059 | 51316.3 | 2887.06 | -5554.06 | -49456 | 0 |
| 2060 | 52990.3 | 2969.98 | -5725.32 | -51039.3 | 0 |

**Supplementary Figure S5: Cumulative change in deaths for both sexes.**

Cumulative difference in deaths

Change from BAU

|  | 1. Maximal activity | 2. Meet guidelines | 3. Revert to 2018 | 4. Everyone inactive |  |
| --- | --- | --- | --- | --- | --- |
| 2021 | -111.01 | -18.8181 | 10.9731 | 83.6191 | 0 |
| 2022 | -448.422 | -83.5893 | 46.5621 | 359.194 | 0 |
| 2023 | -1052.72 | -204.741 | 112.619 | 873.103 | 0 |
| 2024 | -1922.22 | -377.855 | 210.422 | 1625.1 | 0 |
| 2025 | -3030.57 | -593.823 | 338.358 | 2592.49 | 0 |
| 2026 | -4352.45 | -840.319 | 493.241 | 3746.95 | 0 |
| 2027 | -5850.96 | -1108.44 | 672.136 | 5048.52 | 0 |
| 2028 | -7499.32 | -1390.09 | 872.467 | 6475.32 | 0 |
| 2029 | -9275.78 | -1680.32 | 1091.98 | 8005.25 | 0 |
| 2030 | -11154 | -1973.63 | 1328.3 | 9617.2 | 0 |
| 2031 | -13117.2 | -2265.38 | 1580.44 | 11300.1 | 0 |
| 2032 | -15151.9 | -2554.67 | 1846.34 | 13042.1 | 0 |
| 2033 | -17249.3 | -2838.79 | 2125.06 | 14832 | 0 |
| 2034 | -19406.3 | -3117.59 | 2415.7 | 16662.6 | 0 |
| 2035 | -21615.5 | -3391.2 | 2716.73 | 18528.2 | 0 |
| 2036 | -23854.6 | -3655.37 | 3026.08 | 20414.6 | 0 |
| 2037 | -26190 | -3921.99 | 3353.15 | 22388.3 | 0 |
| 2038 | -28608.3 | -4189.46 | 3695.38 | 24438.9 | 0 |
| 2039 | -31101.8 | -4457.41 | 4050.43 | 26558.5 | 0 |
| 2040 | -33660.4 | -4725.45 | 4416.04 | 28738.9 | 0 |
| 2041 | -36245.3 | -4987.79 | 4787.67 | 30956.2 | 0 |
| 2042 | -38858.3 | -5245.67 | 5163.04 | 33208.6 | 0 |
| 2043 | -41497.3 | -5500.17 | 5539.67 | 35493.7 | 0 |
| 2044 | -44144.7 | -5749.83 | 5914.49 | 37790.6 | 0 |
| 2045 | -46790.6 | -5994.18 | 6285.25 | 40092.6 | 0 |
| 2046 | -49425.7 | -6233.44 | 6649.46 | 42394 | 0 |
| 2047 | -52041.7 | -6466.16 | 7005.53 | 44680.7 | 0 |
| 2048 | -54629.8 | -6692.8 | 7352.21 | 46942.4 | 0 |
| 2049 | -57181.8 | -6912.62 | 7688.61 | 49171.5 | 0 |
| 2050 | -59679.7 | -7123.81 | 8012.78 | 51353.8 | 0 |
| 2051 | -62125.1 | -7326.95 | 8324.7 | 53484 | 0 |
| 2052 | -64490.7 | -7518.78 | 8622.21 | 55547.9 | 0 |
| 2053 | -66781.7 | -7701.05 | 8905.82 | 57547.3 | 0 |
| 2054 | -68997.8 | -7873.95 | 9176.7 | 59480.7 | 0 |
| 2055 | -71099.5 | -8031.55 | 9431.31 | 61320.5 | 0 |
| 2056 | -73132.6 | -8182.99 | 9672.96 | 63094.5 | 0 |
| 2057 | -75084.8 | -8326.29 | 9901.72 | 64793.8 | 0 |
| 2058 | -76950.6 | -8460.62 | 10117.6 | 66412.7 | 0 |
| 2059 | -78744.5 | -8588.08 | 10322.3 | 67958.6 | 0 |
| 2060 | -80458.2 | -8708.44 | 10514.5 | 69419.8 | 0 |

**Supplementary Table S6: Comparison of disease groups contributing to overall change in health spending and HALYs in models 1 and 2, over 20 years, 0% discount rate.**

Note: Model 1 contains all pathways to disease and includes injuries due to physical activity. Model 2 contains all pathways to disease, but excludes injuries due to physical activity.

|  |  | All pathways including injury due to activity (model 1) | | | All pathways excluding injury due to activity (model 2) | | |
| --- | --- | --- | --- | --- | --- | --- | --- |
|  |  | **All** | **Female** | **Male** | **All** | **Female** | **Male** |
| **Change in health spending (millions)** | **Total** | -748 | -2,060 | 1,330 | -19,900 | -10,700 | -9,150 |
|  |  | (-6,980 to 4,460) | (-5,900 to 1,010) | (-1,310 to 3,510) | (-25,800,to -15,300) | (-14,500 to -8,040) | (-11,600 to -7,130) |
|  | Cancer and other neoplasms | -1,310 | -862 | -449 | -1,330 | -880 | -444 |
|  |  | (-2,200 to -720) | (-1,420 to -491) | (-844 to -202) | (-2,150 to -700) | (-1,380 to -468) | (-825 to -205) |
|  | Cardiovascular diseases | -2,350 | -726 | -1,620 | -2,390 | -743 | -1,640 |
|  |  | (-4,160 to -1,110) | (-1,420 to -198) | (-2,750 to -855) | (-4,160 to -1,160) | (-1,440 to -226) | (-2,730 to -883) |
|  | Endocrine disorders | -4,400 | -2,010 | -2,390 | -4,370 | -1,990 | -2,370 |
|  |  | (-6,530 to -2,990) | (-2,990 to -1,360) | (-3,540 to -1,630) | (-6,370 to -2,950) | (-2,920 to -1,340) | (-3,440 to -1,610) |
|  | Injuries | 16,900 | 7,230 | 9,690 | -2,250 | -1,450 | -800 |
|  |  | (15,100 to 18,600) | (6,120 to 8,230) | (8,930 to 10,400) | (-3,860 to -957) | (-2,480 to -616) | (-1,390 to -338) |
|  | Kidney and urinary diseases | -714 | -281 | -433 | -707 | -277 | -429 |
|  |  | (-1,250 to -373) | (-493 to -147) | (-753 to -227) | (-1,260 to -395) | (-498 to -154) | (-762 to -240) |
|  | Mental and substance use disorders | -7,960 | -4,970 | -2,980 | -7,820 | -4,880 | -2,940 |
|  |  | (-13,500 to -4,270) | (-8,440 to -2,640) | (-5,060 to -1,610) | (-13,600 to -4,110) | (-8,530 to -2,550) | (-5,050 to -1,570) |
|  | Neurological conditions | -580 | -284 | -296 | -566 | -277 | -290 |
|  |  | (-1,330 to -133) | (-660 to -61.4) | (-672 to -71.5) | (-1,280 to -102) | (-630 to -46) | (-645 to -60.1) |
| **Change in HALY (thousands)** | **Total** | 653 | 450 | 202 | 1,520 | 874 | 647 |
|  |  | (230 to 1,210) | (192 to 790) | (37 to 418) | (1,100 to 2,060) | (616 to 1,210) | (487 to 860) |
|  | Cancer and other neoplasms | 44.5 | 26.2 | 18.5 | 45 | 26 | 18 |
|  |  | (25 to 75.1) | (15.3 to 41.7) | (8.7 to 33.8) | (24.1 to 73.3) | (14.8 to 41.2) | (8.6 to 32.8) |
|  | Cardiovascular diseases | 191 | 90.5 | 100 | 190 | 90 | 100 |
|  |  | (135 to 260) | (64.3 to 124) | 100 (71.1 to 137) | (133 to 267) | (62.2 to 126) | (70.3 to 141) |
|  | Endocrine disorders | 239 | 116 | 122 | 239 | 116 | 122 |
|  |  | (171 to 329) | (82.9 to 161) | (88.2 to 168) | (172 to 323) | (83.1 to 158) | (88.4 to 165) |
|  | Hearing and vision disorders | 0.44 | 0.23 | 0.2 | 0.42 | 0.23 | 0.19 |
|  |  | (0.03 to 1.05) | (0.02 to 0.57) | (0.01 to 0.48) | (0.05 to 1.04) | (0.03 to 0.56) | (0.03 to 0.48) |
|  | Injuries | -761 | -360 | -400 | 116 | 69 | 48 |
|  |  | (-884 to -654) | -360 (-426 to -302) | (-458 to -352) | (49.8 to 202) | (29.5 to 120) | (20.3 to 82.8) |
|  | Kidney and urinary diseases | 14.5 | 6.8 | 7.66 | 15 | 7 | 8 |
|  |  | (7.54 to 25.7) | (3.58 to 12.1) | (4.01 to 13.6) | (8.01 to 25.5) | (3.77 to 12) | (4.25 to 13.5) |
|  | Mental and substance use disorders | 866 | 534 | 333 | 857 | 528 | 329 |
|  |  | (485 to 1,390) | (298 to 861) | (186 to 531) | (459 to 1,390) | (282 to 856) | (174 to 530) |
|  | Neurological conditions | 44.5 | 28.5 | 16 | 44 | 28 | 16 |
|  |  | (10.5 to 98.5) | (6.44 to 63.6) | 16 (4 to 34.8) | (8.79 to 101) | (5.41 to 65) | (3.32 to 35.7) |

**Supplementary Table S6: Main model outputs for scenarios 1-4 over 20 years, 3% discount rate.**

| Metric | Sex | Baseline | Everyone at 4200+ MET/min/week | All sedentary to 600-1200 MET/min/week | Revert to 2018 physical activity level | No physical activity |
| --- | --- | --- | --- | --- | --- | --- |
| **Total HALY (thousands)** | All | 306,000 | 443 (126 to 859) | 156 (95 to 243) | -108 (-178 to -58) | -941 (-1,590 to -478) |
|  | Female | 153,000 | 315 (121 to 573) | 87.6 (52.6 to 137) | -66.6 (-110 to -36.2) | -503 (-839 to -270) |
|  | Male | 153,000 | 128 (5.41 to 291) | 68.4 (41.7 to 105) | -41.4 (-68.5 to -21.8) | -438 (-758 to -210) |
| **Life years gained (thousands)** | All | 365,000 | 169 (127 to 226) | 27.3 (15.6 to 42.2) | -21 (-29.4 to -14.9) | -145 (-214 to -94.2) |
|  | Female | 185,000 | 84.7 (63 to 113) | 13.8 (7.74 to 21.5) | -11.1 (-15.6 to -7.7) | -56.5 (-82.6 to -36.9) |
|  | Male | 179,000 | 84.9 (63.2 to 113) | 13.5 (7.92 to 20.7) | -9.96 (-14 to -7.08) | -88.8 (-131 to -58) |
| **Total health spending (millions)** | All | 2,700,000 | 70.9 (-4,620 to 3,880) | -1,400 (-2,350 to -684) | 1,010 (424 to 1,780) | 6,850 (2,010 to 13,400) |
|  | Female | 1,420,000 | -1,270 (-4,150 to 1,000) | -768 (-1,310 to -368) | 582 (236 to 1,040) | 4,240 (1,870 to 7,580) |
|  | Male | 1,280,000 | 1,350 (-603 to 2,950) | -631 (-1,060 to -300) | 422 (167 to 758) | 2,630 (111 to 5,960) |
| **Total income (millions)** | All | 8,960,000 | 11,600 (8,780 to 15,300) | 779 (498 to 1,170) | -1,370 (-1,850 to -1,010) | -11,700 (-16,100 to -8,290) |
|  | Female | 3,590,000 | 6,240 (4,740 to 8,290) | 358 (232 to 529) | -743 (-999 to -552) | -4,780 (-6,590 to -3,420) |
|  | Male | 5,370,000 | 5,300 (3,990 to 6,960) | 423 (252 to 670) | -627 (-855 to -447) | -6,890 (-9,610 to -4,770) |
| **Total deaths** | All | 3,210,000 | -23,700 (-31,000 to -18,000) | -3,440 (-5,290 to -2,010) | 3,080 (2,210 to 4,230) | 20,300 (13,400 to 29,300) |
|  | Female | 1,550,000 | -11,900 (-15,500 to -9,010) | -1,730 (-2,680 to -991) | 1,580 (1,130 to 2,170) | 7,820 (5,230 to 11,200) |
|  | Male | 1,670,000 | -11,900 (-15,600 to -9,010) | -1,710 (-2,640 to -1,010) | 1,500 (1,070 to 2,070) | 12,500 (8,260 to 18,100) |
| **Deaths before age 75** | All | 782,000 | -6,770 (-8,880 to -5,140) | -659 (-1,090 to -377) | 1,120 (805 to 1,580) | 8,500 (5,700 to 12,600) |
|  | Female | 302,000 | -2,460 (-3,170 to -1,910) | -200 (-323 to -119) | 389 (285 to 534) | 2,230 (1,540 to 3,190) |
|  | Male | 480,000 | -4,310 (-5,760 to -3,230) | -458 (-775 to -252) | 735 (516 to 1,050) | 6,270 (4,110 to 9,370) |
